# The INDSCI-SIM model for COVID-19 in India

**DOI:** 10.1101/2021.06.02.21258203

**Authors:** Dhiraj Kumar Hazra, Bhalchandra S. Pujari, Snehal M. Shekatkar, Farhina Mozaffer, Sitabhra Sinha, Vishwesha Guttal, Pinaki Chaudhuri, Gautam I. Menon

## Abstract

Estimating the burden of COVID-19 in India is difficult because the extent to which cases and deaths have been undercounted is hard to assess. The INDSCI-SIM model is a 9-component, age-stratified, contact-structured compartmental model for COVID-19 spread in India. We use INDSCI-SIM, together with Bayesian methods, to obtain optimal fits to reported cases and deaths across the span of the first wave of the Indian pandemic, over the period Jan 30, 2020 to Feb 15, 2021. We account for lock-downs and other non-pharmaceutical interventions, an overall increase in testing as a function of time, the under-counting of cases and deaths, and a range of age-specific infection-fatality ratios. We first use our model to describe data from all individual districts of the state of Karnataka, benchmarking our calculations using data from serological surveys. We then extend this approach to aggregated data for Karnataka state. We model the progress of the pandemic across the cities of Delhi, Mumbai, Pune, Bengaluru and Chennai, and then for India as a whole. We estimate that deaths were undercounted by a factor between 2 and 5 across the span of the first wave, converging on 2.2 as a representative multiplier that accounts for the urban-rural gradient across the country. We also estimate an overall under-counting of cases by a factor of between 20 and 25 towards the end of the first wave. Our estimates of the infection fatality ratio (IFR) are in the range 0.05 - 0.15, broadly consistent with previous estimates but substantially lower than values that have been estimated for other LMIC countries. We find that approximately 40% of India had been infected overall by the end of the first wave, results broadly consistent with those from serosurveys. These results contribute to the understanding of the long-term trajectory of COVID-19 in India.

## Introduction

COVID-19, a disease of zoonotic origin whose causative agent is the beta coronavirus SARS-CoV-2, is believed to have first infected humans towards the latter part of November 2019, in or near the Chinese city of Wuhan [1]. It then spread, aided by international travel networks, around the world, with devastating epidemics in the USA, the UK, Europe and South America, even as cases in China declined [2]. With close to 170 million recorded cases and 3.5 million recorded deaths worldwide as of the end of May 2021, the COVID-19 pandemic can be expected to be the most consequential epidemiological event of our lifetimes.

The first COVID-19 case in India was detected at the end of January, 2020 [3]. By March 25, 2020, the total numbers of Indian cases had increased to just above 600. At that point, the Indian government ordered a sequence of stringent country-wide lockdowns that were to last for 68 days (March 25 - June 1, 2020) [4]. The lockdown was then relaxed in multiple phases. All through, reported cases of COVID-19 kept rising, although the pace of increase was arguably held in check by the stringency of the lockdown [5]. A peak of about 98,000 cases was reached in mid-September. Cases in India then declined steadily for the next four months, even as cases rose elsewhere in the world. By mid-January, it appeared as if India might have avoided the multiple waves of cases seen elsewhere. However, numbers at the level of individual cities and states presented a more complex story. Delhi and Mumbai, for example, saw multiple waves of cases [6]. The decline in cases at the all-India level persisted from the peak around mid-September to the middle of February, when they began to increase again [7]. This increase has been linked to the emergence of more transmissible variants, specifically the B.1.617 and B.1.1.7 variants, as well as to a relaxation of COVID-appropriate behaviour [8]. The pace of this increase was far steeper than the pace at which the first wave of cases were recorded. (As of roughly mid-May 2021, the decline of the second wave, from a peak of a little more than 410,000 cases daily at its maximum, has begun.) A time-line of the first wave of COVID-19 in India is displayed in Fig. 1. This figure shows the increase in cumulative cases, in tests and in deaths, highlighting the period of the national lockdown.

**Fig 1.**
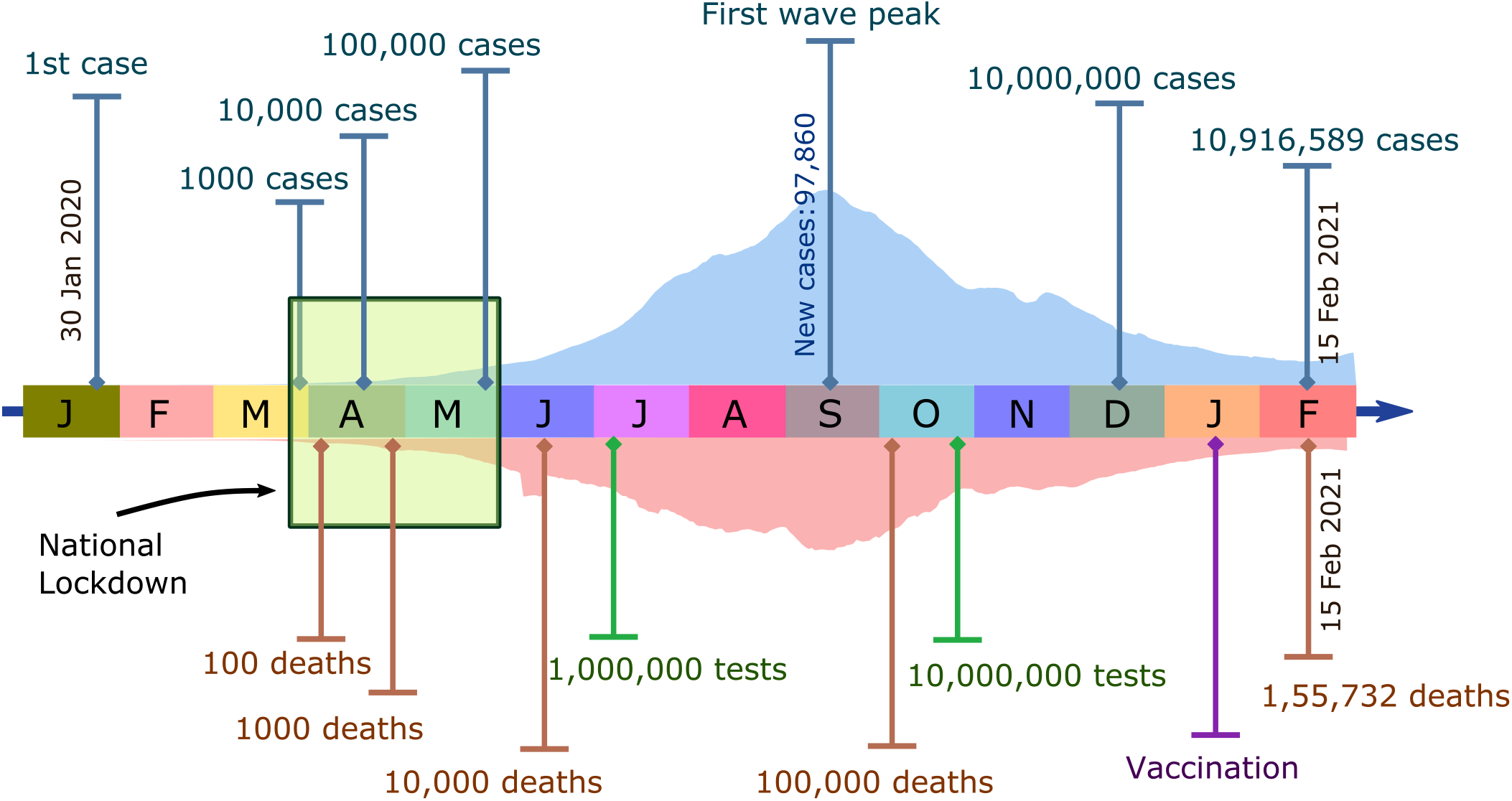
The timeline of the first wave of COVID-19 in India, beginning from the time of the first detected cases in India and ending on February 15, 2021, across different months during that period. The period of India’s nationwide lockdown, between the dates of March 25 and May 31, 2020, is shown as a green box. Daily infection and death curves are shown in filled light blue curve and light brown inverted curves respectively. Specific milestone values for cases, deaths and tests are also provided.

Epidemiological models are useful because they allow us to reason about parameters that control pandemic spread, and how interventions serve to modulate them, extrapolating from a trajectory of cases and deaths. Population-level epidemiological information, such as results from serological surveys, help to further constrain these models. The earliest models for COVID-19 in India come from the work of Mandal *et. al*. [9]. This compartmental model addressed two issues, the effects of imperfect airport screening measures and the question of optimal strategies for mitigation once the disease had spread to the major Indian cities. Chatterjee et al. used a stochastic SEIR model to examine the effects of lockdowns on case counts [10]. Work from the group of Bhramar Mukherjee provided early insights into the progress of the pandemic and continues to do so [11]. Their work uses a Bayesian extension of the SIR model, the extended susceptible-infected-removed (eSIR) model, to project case-counts and deaths. Agent based models have provided useful insights, at the level of full cities, into mitigation methods and the effectiveness of non-pharmaceutical interventions [12]. Related references which model COVID-19 in India are [10, 13–23, 23–36]. These models are very largely compartmental models of varying degrees of complexity [37]. Almost all of them were aimed at understanding the initial stages of the evolution of the pandemic and the role of interventions. To our knowledge, virtually none of them, with the exception of Ref. [11], have described the full trajectory of the epidemic.

As the second most populous nation in the world, with a population of close to 1.4 billion, the consequences of an explosion of COVID-19 cases in India could easily dwarf its impact anywhere else [38]. What remains unclear is the extent to which the Indian population has so far been infected by COVID-19 and whether any proximity to herd immunity through infection might slow later waves of disease [39–41]. Large-scale serological surveys (serosurveys) from the Indian Council of Medical Research (ICMR), adjusted for test sensitivity, estimate the overall fraction of those with a prior COVID-19 infection to be about 22% by December 2020-January 2021 [42–44]. The first two ICMR serosurveys obtained a nation-wide seroprevalence of 0.73% in May-June 2020 and of 6.6% in August-September 2020. A strong gradient of seroprevalance between urban and rural India has been a consistent feature of these national serosurveys. These were done in just 70 districts of a total of about 740 in India, however, so only represent a relatively small cross-section of the country. Other serosurveys have studied specific Indian cities, among them Bengaluru [41], Chennai [45, 46], Delhi [47], Pune [48] and Mumbai [49]. These city-based surveys estimate that fairly substantial fractions of the urban population should have been infected by the time that the first wave waned. In many cases, such as for Delhi, Mumbai and Pune, this fraction has been estimated at over 50%. However, because of the variety of test kits used, it has proved hard to compare results from different serosurveys. Also, these presumed high levels of prior seropositivity seem to have done little to offset the dramatic rise of cases seen at the onset to the second wave in urban India. This raises questions of potential errors in the serosurveys tied to the sensitivity and specificity of the kits used, as well as of the importance of reinfections [50].

This paper describes INDSCI-SIM, a age-stratified, contact-structured compartmental model for COVID-19 spread in India. INDSCI-SIM is a 9-component model, adopted and modified from [51], with rates bench-marked to a wide range of available data. It projects numbers of both mild and severe cases and can be generalized to a variety of India-specific situations. Finally, it can be used to incorporate the modelled effects of a number of public health interventions, including lock-downs as well as progressive improvements in case identification. At the methodological level, our techniques can account for improvements, with time, in case identification as well as in treatment leading to lower overall mortality, within a fully Bayesian framework.

Our work describes the trajectory of COVID-19, including fits to both cases and deaths, across all districts in the southern Indian state of Karnataka, as well as in the capital of that state, Bengaluru. Also included are similar fits to cases and deaths in multiple Indian cities, including Mumbai, Delhi, Chennai and Pune, as well as to aggregate data for the whole country. Via this excersise, we estimate that approximately 40-50% of India was infected at the time the second wave struck, roughly consistent with serosurvey results. We suggest that cases have been under-counted by a factor ranging from about 90 at the onset of the Indian epidemic to about 20 at the end of the first wave. We estimate that a multiplicative factor of about 2.2 between counted and actual deaths might be a reasonable estimate across India for the first wave of the pandemic, although even this estimate relies on a number of approximations. Finally, our results are consistent with the observation that the trajectory of the disease across India has been inhomogeneous, with complex spatio-temporal behaviour at the level of districts and states summing to give smoother results for the country as a whole.

## 1 Materials and Methods

The epidemiological compartmental model represented by INDSCI-SIM is shown in Fig. 2. INDSCI-SIM is based on a model introduced in Ref. [51], that expands the classical SIR & SEIR framework with compartments that account for an asymptomatic infectious state [51–53]. The model also accounts for variations in the severity of disease across the infected class [54]. There is a compartment for hospitalized cases as well as compartments that count deaths as well as recoveries. The model parametrizes the transmission of infection from the infected classes on the susceptible class, also allowing this to depend on time. This is an indirect, yet useful, way of describe increased stringency as well as relaxations in non-pharmaceutical interventions. These include both direct effects such as increased testing, the imposition of mask-wearing and the effects of isolation and quarantines. They also include, *inter alia*, indirect effects, such as increased public awareness and related modification of behaviour [55].

**Fig 2.**
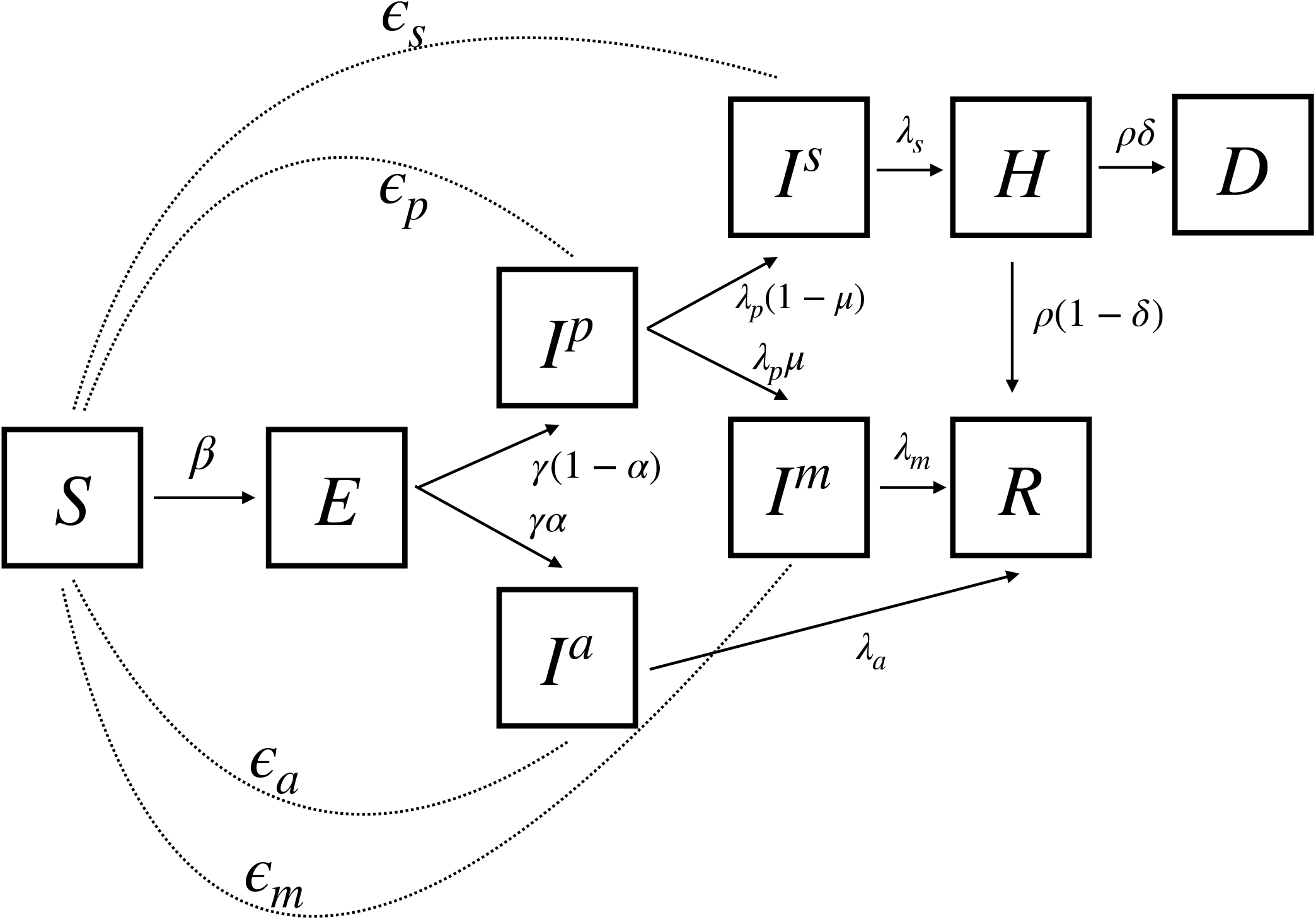
Schematic diagram of the compartmental model used in this analysis, adopted from [51]. The dotted lines indicate the force of infection from the infected compartments on the susceptible population. Transitions between compartments, denoted via solid lines and an arrow, are defined as in Equation 1 which contains both bare rates as well as information as to how flow is to be divided between compartments. The number of persons transiting from one compartment to the other in each day depends on the product of the transition rates and the branching fractions.

### 1.1 Disease Progression

SARS-CoV-2 infection manifests as an acute respiratory infection, progressing to respiratory failure in a small number of patients [56–58]. It can result in a range of clinical manifestations, from asymptomatic or mild infection to severe, requiring hospitalization [59]. Among patients who are symptomatic, the median incubation period is approximately 4 to 5 days [60]. About 97% have symptoms within 11 days after infection [61]. These can further be categorized as mild, severe and critical [62]. Patients who are hospitalized, the severe category, can progress to severe pneumonia and acute respiratory distress syndrome (ARDS). A fraction of these patients will require ventilation and an even smaller fraction may die [63]. Older patients experience greater clinical severity of COVID-19, which we account for via age-stratified parameters [63, 64]. Co-morbidities such as cardiovascular disease, diabetes and obesity are common underlying conditions associated with worse clinical outcomes and increased disease severity [64–66]. These can be incorporated through a composite risk score affecting branching rates between mild and severe disease states, although we do not do so here. Males may experience more severe disease than females, and genetic variations, including the ABO blood type, have been implicated in clinical outcomes for patients with COVID-19. Our model ignores these effects [67, 68]. Around 40–75% of infections may be asymptomatic, a fairly broad range. We note that numbers for India suggest a larger fraction of asymptomatic cases than reported elsewhere [69, 70].

### 1.2 Transmission dynamics

The compartmental structure we consider contains susceptible (*S*), exposed (*E*), asymptomatic infectious (*I*^*a*^), pre-symptomatic infectious (*I*^*p*^), mildly symptomatic infectious (*I*^*m*^), severely symptomatic infectious (*I*^*s*^), hospitalized (*H*), dead (*D*) and recovered (*R*) compartments. Transitions between these model compartments are shown in Figure 2. The dotted lines indicate the force of infection from the infected compartments on the susceptible population. Frequency-dependent transmission is assumed. As appropriate for a fast-spreading infection, we ignore the effects of demography [52].

The model equations, for unstructured compartments, are

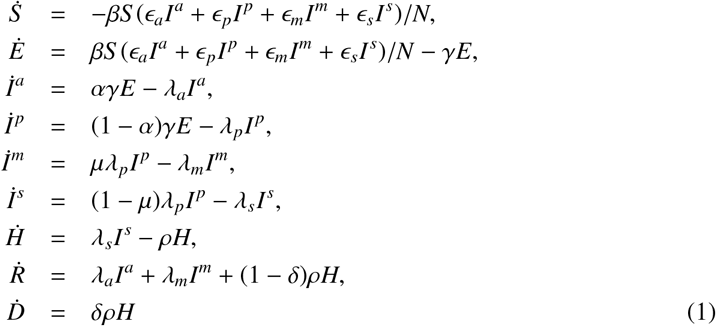

The left-hand side of these equations denote first-order time derivatives. The population size is *N*. Infectious individuals in any associated compartment can infect the susceptible population regardless of symptoms and severity with a fixed transmission rate *β*. However, this quantity is modulated by the relative intensity of contacts between susceptible and infectious individuals whose effect is simply specified here through factors of *ϵ*. These can, in principle, vary with time and can also be chosen to vary between compartments.

Each infected individual enters an ‘exposed’ compartment (*E*), spending an average latent period 1*/γ* days before becoming infectious [71]. A fraction *α* of this infectious population remains asymptomatic (*I*^*a*^) until recovery. Asymptomatic individuals are assumed to be infectious for an average period of 1*/λ*_*a*_ days [72, 73]. Those in the remaining fraction, of (1 − *α*), enter a pre-symptomatic state (*I*^*p*^) where clinical symptoms are not exhibited for a relatively short average period of 1*/λ*_*p*_ [72]. Individuals in this pre-symptomatic compartment go on to developing mild or severe symptoms [74]. All rates applicable to the model are provided in Table 1.

**Table 1.**
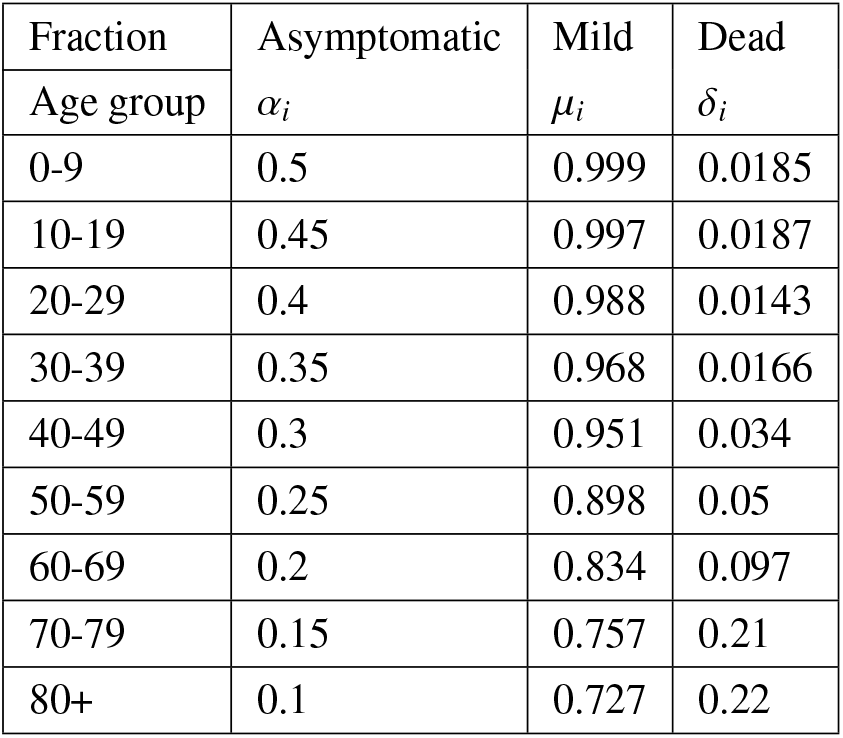
Age dependent branching ratios between compartments. Our values for *α*_*i*_ and *µ*_*i*_ are from those consolidated by the COVASIM program, Refs. [78] and [79]. We use IFR’s from data from LMIC’s taken from fits in the paper of Ref. [80], where the following formula is derived: *log*_10_(I*FR*) = − 3.27 + 0.0524 * a*ge*. We modify only the *δ*_*i*_ for the 80+ age group to account for the leveling off of mortality in older age groups described by Ref. [25]. The IFR for each age group can be obtained as IFR_*i*_ = (1 − *α*_*i*_)(1 − *µ*_*i*_)*δ*_*i*_. We use the estimated *δ*_*i*_ numbers as our initial IFRs, although when we allow them to vary as part of our minimization strategy to find the effective IFR’s we multiply all *δ*_*i*_’s by a single smooth time-varying factor, optimizing this against data.

| Fraction | Asymptomatic | Mild | Dead |
| --- | --- | --- | --- |
| Age group | $\alpha_i$ | $\mu_i$ | $\delta_i$ |
| 0-9 | 0.5 | 0.999 | 0.0185 |
| 10-19 | 0.45 | 0.997 | 0.0187 |
| 20-29 | 0.4 | 0.988 | 0.0143 |
| 30-39 | 0.35 | 0.968 | 0.0166 |
| 40-49 | 0.3 | 0.951 | 0.034 |
| 50-59 | 0.25 | 0.898 | 0.05 |
| 60-69 | 0.2 | 0.834 | 0.097 |
| 70-79 | 0.15 | 0.757 | 0.21 |
| 80+ | 0.1 | 0.727 | 0.22 |

*A fraction µ* of symptomatic cases is assumed to develop mild symptoms (*I*^*m*^), while the remaining fraction (1 − *µ*) of cases are transferred to the severe class (*I*^*s*^). Infectious cases with mild symptoms recover without hospitalization after 1*/λ*_*m*_ days. Severe cases require hospitalization after an average of 1*/λ*_*s*_ days. From the hospitalized population (*H*), we assume that a proportion (1 − *δ*) recovers successfully (*R*), after spending an average duration of hospitalization 1*/ρ* days. The remaining fraction, *δ*, of the hospitalised, will die. The numerical values of these parameters are specified in Table 1 [75, 76].

Of these parameters, the infectivity parameter *β* is particularly central. It determines the effective reproduction ratio as the epidemic proceeds. The force of infection arising from asymptomatic cases alone is assumed to be lower in comparison to that arising from the pre-symptomatic, mildly symptomatic and severely symptomatic cases.

#### 1.2.1 Age-structured model

We incorporate age-structuring, dividing each compartment into sub-compartments, representing the age-intervals 0 − 9, 10 − 19, 20 − 29, 30 − 39, 40 − 49, 50 − 59, 60 − 69, 70 − 79 and 80+. We denote the sub-compartment by adding the subscript *i*, for each age-bracket, to each of the compartment labels, thus obtaining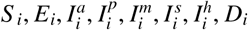. We define *β*_*ij*_ as the bare infectivity term coupling age-brackets *i* and *j*, although we will assume *β*_*ij*_ ≡ *β* here. The quantities 𝒞_*ij*_ defines the intensity of contacts between these age-brackets. We assume that, during the lockdown, only contacts at home are effective, since schools and workplaces are closed. We can also assume that contacts in public transports and other places are negligible. Since, in India, schools largely remained closed even after the lockdown ended and workplace crowding had reduced substantially, we assume that even without any lockdown, work contacts are only 50% effective. These contacts are obtained from those compiled by Ref. [77]. The appropriate contact matrices are discussed in the SI (Ref. SI: Section 2, Figure 1.).

To incorporate time-dependence in contacts, we generalize the four *ϵ* terms, *ϵ*_*a*_, *ϵ*_*s*_ and *ϵ*_*m*_ and *ϵ*_*p*_, making their magnitude time-dependent. This allows for varying implementation of testing, quarantining and isolation rules, lock-downs and other non-pharmaceutical interventions.

Our generalized equations, for *N*_*a*_ age-brackets, are then

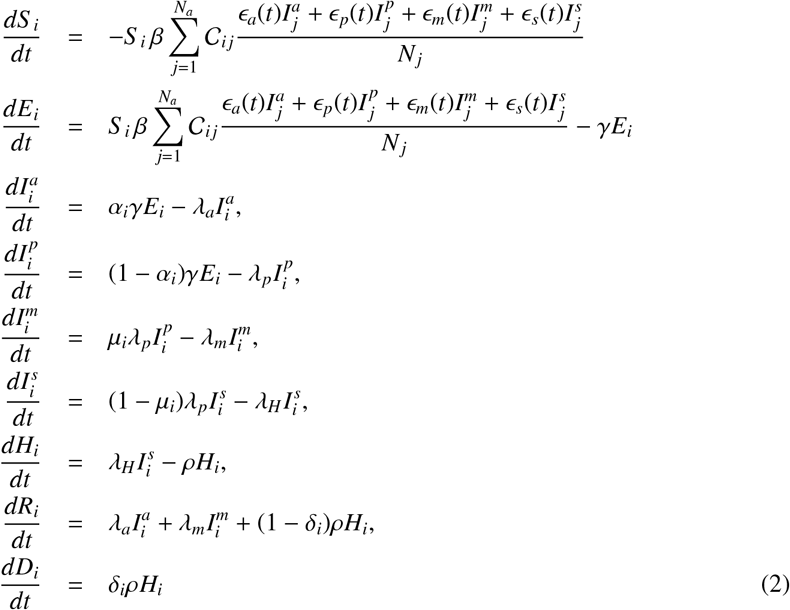

The values used here are listed in Table 1

### 1.3 Time-dependence of effective contacts to model non-pharmaceutical interventions

The effective infectivity in the INDSCI-SIM model is a product of three terms. The first is a bare infectivity parameter (*β*_*ij*_ ≡ *β*)and the second is the term involving the contacts, the *C*_*ij*_’s. The third is the temporal modulation, the factors of *ϵ*. While it is only variation in this overall product that is of significance, it allows us to conceptualize interventions in a more targeted way: the sudden changes in infectivity imposed by a lockdown can be associated with abrupt changes in *β* whereas slow and secular improvements in masking etc. can be associated to the smoothly varying factors of *ϵ*. This product is affected by mask-wearing, hand-washing, voluntary self-isolation or self-quarantining and the maintenance of social-distancing. It is also influenced by global non-pharmaceutical interventions such as closures of schools, malls and cinemas, large-scale lock-downs, restrictions on movement and autonomous modifications of social behaviour.

We first use a “global” value of *β*, one that differs in the lockdown period and in the open period. We then model compliance with restrictions by modulating this with a time-dependent factor, using a hyperbolic tangent function of time, as described below. This function comes with an associated time-scale which is an output of our minimization procedure. A quantification of mask-wearing India-wide comes from studies incorporated into the IHME model for India that are standardized using survey results from the University of Maryland Social Data Science Center, from the Kaiser Family Foundation and the YouGov COVID-19 Behaviour Tracker survey [81]. This data motivates the use of such an interpolating function, since it shows an initial lag period, a sharp rise as public awareness increases and saturation at about 70% reflecting social acceptance of mask-wearing.

We also allow for a decay of the relative intensity of contacts between susceptible and infected individuals by assuming an exponential decay of the *ϵ*_*i*_ terms, viz.,

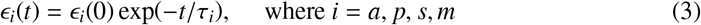

Here, *τ*_*i*_ represents the characteristic time-scale describing the increased effectiveness of non-pharmaceutical interventions (NPIs). These could include restrictions on crowding, improvements in screening procedures as well as increased testing. The net effect of these could be chosen to be different for each of the infectious categories. Although the force-of-infection ultimately involves a product of both the *β*(*t*) and the *ϵ* (*t*) terms, splitting them out thus provides somewhat more control over the specifics of the non-pharmaceutical interventions as well as the ability to account for new variants that might affect the value of *β* but not the *ϵ*-s.

### 1.4 Estimating *R*_0_

Given the central equations defining our model, we compute the dominant eigenvalue of the next generation matrix [82] obtained from Equation 1. This is the basic reproductive ratio, *R*_0_. This result, derived using a next-generation method outlined in SI, yields

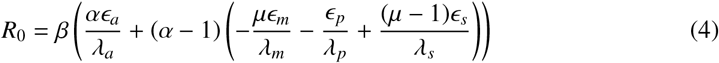

Note that in Equation 1 the relative intensity of the contacts (*ϵ*) are simply constants.

### 1.5 Estimating *R*(*t*)

The time evolution of *R*(*t*) is obtained from Equation 2. We obtain the next generation matrix (ℱ 𝒱^−1^), incorporating age stratification and contract matrices. Lockdown changes the contact matrix; we take this into account in our estimation of *R*(*t*). The computation of the next generation matrix, and thence *R*(*t*) corresponding to Equation 2, is discussed in SI: Section 1. Comparing model predictions with the data generates samples of parameters. From these samples we compute the bounds on *R*(*t*) for each region of interest.

### 1.6 Computing effective *R*(*t*) from the data

A standard way to describe infection spread is to calculate the effective reproduction rate *R*(*t*) at a given time *t*. To estimate *R*(*t*) we use a Bayesian approach developed by Bettencourt and Ribeiro [83], later modified by K. Systrom [84]. In this approach, given *k* new cases, the probability distribution of *R*(*t*) on a certain day *t* is:

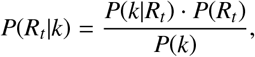

where *P* (*k* | *R*_*t*_) is the likelihood of seeing *k* new cases given *R*(*t*) (this is assumed to follow a Poisson distribution), *P*(*R*_*t*_) is the prior, and *P*(*k*) is the probability of seeing *k* cases. The method of deciding the appropriate priors is described in Ref. [83, 84].

### 1.7 Fixed Parameters

Several parameters determine the progress of the pandemic in the INDSCI-SIM model. These include parameters which remain fixed, such as the rate of transitions between the compartments, described above, and a choice for the initial IFR. Other parameters are allowed to vary so as to model the effects of non-pharmaceutical interventions or optimized to fit available data, as discussed below. We assume that asymptomatic patients are less likely to pass on infection to susceptibles by a factor of 2/3, consistent with data from Ref. [69, 85]. The choices of our fixed parameters are provided in Table 2.

**Table 2.** The transition rates between compartments and the efficiency parameters. These parameters are fixed during the analysis and their values are taken from [51, 86]

| Parameter | Rate (1/day) | Description |
| --- | --- | --- |
| $\gamma$ | 0.5 | Transition rate from exposed to asymptomatic or pre-symptomatic (mean 2 days) |
| $\lambda_a$ | 0.1428 | Transition rate from asymptomatic to recovered (mean 7 days) |
| $\lambda_m$ | 0.1428 | Transition rate from mild to recovered (mean 7 days) |
| $\lambda_p$ | 0.5 | Transition rate from pre-symptomatic to mild and severe (mean 2 days) |
| $\lambda_s$ | 0.1736 | Transition rate from severe to hospitalized (mean 6 days) |
| $\rho$ | 0.068 | Transition rate from hospitalized to recovered or dead (mean 15 days) |
| Parameter | Efficiency |  |
| $\epsilon_a$ | 0.67 | Relative intensity of contacts for asymptomatic |
| $\epsilon_p$ | 1 | Relative intensity of contacts for pre-symptomatic |
| $\epsilon_m$ | 1 | Relative intensity of contacts for mild |
| $\epsilon_s$ | 1 | Relative intensity of contacts for severe |

### 1.8 Variable Parameters

We maximize a likelihood function to obtain optimal fits to the data, sampling a broad prior distribution in parameter space to assess uncertainties in our description of the disease progression. Our simulations are initiated around 2 weeks prior to the first death being reported. Since the time evolution of any similar compartmental model with constant parameters and no births or deaths would yield only a single wave, we use an adaptive parametrization to address multiple waves of COVID-19 cases. The parameters that enter our description are described below.

We choose a generic functional form for all our parametrizations that involve parameter changes with time. This form interpolates between a high and a low value, and is a function of a characteristic time-scale. We choose a hyperbolic tangent function for concreteness, and because it is easy to specify, but any other suitable function could be used in its place.

a. Initial exposed (*E*_*initial*_): The exposed people on the first day of simulation, distributed between age groups according to the population fraction.
b. IFR-associated parameters (Δ_IFR_, *δ*^*i*^(*t*_*initial*_), *δ*^*i*^(*t* _*final*_) and *D*_IFR_): A number of studies [87–89], suggest that the IFR has decreased over time. This is attributed to an improved clinical handling, new pharmacological treatments such as corticosteroids, non-pharmacological treatments such as proning and simply earlier interventions and finally the potential prophylactic consequences of lower viral load exposure from masking [87]. For concreteness, we choose a specific functional form to describe this decrease, accounting for it by assuming a smooth transition governed by a hyperbolic tangent function. We model the variation of 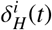 as:

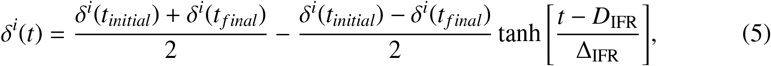

where *D*_IFR_ is the transition time-point (here, tied to the Hospitalized Fatality Ratio) and Δ_IFR_ is the characteristic time width of the transition. This introduces four parameters, *δ*^*i*^(*t*_*initial*_), *δ*^*i*^(*t* _*final*_), the timescale Δ_IFR_ and the transition point itself, *D*_IFR_. Although this formulation is general, allowing for age-bracket dependent variation of the IFRs with time, we choose to allow all age-brackets to have the same functional behaviour, rendering the index *i* redundant. We take *δ*^*i*^(*t* _*final*_) to be a sixth of *δ*^*i*^(*t*_*initial*_): given an initial IFR of 0.3% this potentially allows the IFR to decay across the range 0.3% to 0.05%, over a timescale defined by Δ_IFR_, with the cross-over point between the upper and lower limit defined by *D*_IFR_.
c. Bias(*b*) and bias-variation (Δ_*b*_) : A large fraction of the population remains asymptomatic to COVID-19 infections. In addition, some symptomatic patients may also opt not to be tested. Since testing in India has been limited throughout the first wave, a substantial percentage of actual infections can be expected to have remained undetected, with detected infections always represent an under-counting of the true numbers of infected [90]. We use a bias parameter *b* to accommodate this scaling relation, introducing another parameter to estimate. We parametrise the bias factor as:

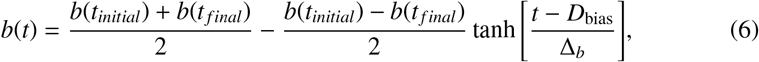

where Δ_*b*_ represents the bias variation timescale. We use the transition time *D*_bias_ to be 3 months from the beginning of March but checked that the results should depend minimally on *D*_bias_ if the priors on the *b*(*t*_*initial*_) and Δ_*b*_ are wide enough. This introduces three parameters *b*(*t*_*initial*_), *b*(*t* _*final*_) and Δ_*b*_. We fix *b*(*t* _*final*_) so that it will reach 1 asymptotically. Our calculation obtains the daily number of infected people from the day. We then fix a multiplier that relates the actual numbers of infected to those detected each day. This number, for those expected to be detected in each day, is then compared with the numbers of those reported infected.

If the reported numbers of daily infected cases, or the daily numbers of deaths, shows multiple peaks, a single parametrization will not capture this behaviour. The sources of this behavior are complex functions of government policy constraining the contacts between people, of the sum of individual actions taken to prevent infection and also of the entry of new potentially more infectious variants. Our analysis must be flexible enough to account for these. Set against this is the requirement that we should not over-determine the model, by allowing for a large number of such changes.

We allow our parametrization to vary in the following way: We assume that the time over which we choose to model the data is divided into *N* segments, with each segment denoted by *i*. This defines *N* − 1 break points or nodes. The infectivity parameter *β* and the timescale *τ* can change in these segments. For simplicity, *τ* is assumed to be the same between segments. We thus parametrize the model with the following:

a. *β*_*i*_: Infectivity within the *i*’th window.
b. *τ*_*i*_: Timescale within the *i*’th window.
c. *Node*_*i*_: The position of the break points (dates) across which *β*_*i*_ and *τ*_*i*_ are change. We decide the required number of break points by comparing the Bayesian evidence for the models with different break points.

Comparisons of the Bayesian evidence indicates that different *τ*_*i*_’s defined in each segment are not favored. Thus, we work with *N β*’s, a single *τ* and *N* − 1 *Node* parameters. For *N* windows, then, we will have *N* + 1 + (*N* − 1) parameters specific to adaptive parametrization, 4 parameters (*E*_*initial*_, *D*_IFR_, Δ_IFR_, *b*, Δ_*b*_) common to both parametrizations and 2 noise error parameters (for modelling the fluctuations in the infected and death data discussed in subsubsection 1.9.1).

### 1.9 Bayesian modeling

Our model is a mechanistic model that should capture the significant aspects of the dynamics of COVID-19 disease spread. It has a number of parameters which must be optimized. We achieve this optimization through a simultaneous fit to the data on deaths and on detected cases. The latter is related to the “true” number of cases through the time-dependent bias factor.

We address this optimization through Bayesian methods. In such methods, probability distributions over parameters, and not point estimates, are obtained [91, 92]. Bayesian models require a prior distribution over parameters to be specified as well as a likelihood of observing the data under a specific assignment of parameters. Given parameters and initial conditions, a compartmental model defines a unique solution for each of the compartments.

Such methods begin with prior estimates for the parameters entering the model, usually chosen so that they are at least partially constrained by prior knowledge [91]. A likelihood function, chosen appropriately, estimates the probability with which the observed data can be accounted for by a specific parameter set. This leads to a posterior distribution over the parameter set. Credible (confidence) intervals can be derived from such calculations.

#### 1.9.1 Likelihood Model

We compute the likelihood after a logarithmic transformation, optimizing the product of the likelihoods from the infected and death data as (ℒ | _*infection*_ × ℒ |_*death*_). We choose a form for the likelihood function that accounts for two sources of error [93]. We assume no correlation between the variances of infected and death data and treat them independently in the joint fit. We use a Gaussian log likelihood defined as:

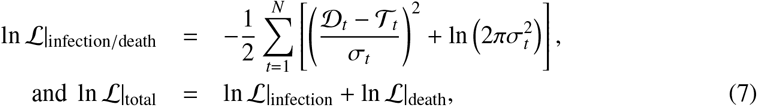

where 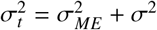 represents the variance in the data. This is the sum of a measurement error *σ*_*ME*_ and of an intrinsic scatter *σ*. 𝒯_*t*_ represents a suitable function (see below) of the computed value of infected/death cases, while 𝒟_*t*_ represents reported infected/death data on day *t* scaled with the same function. The data must first be transformed so that it is normally distributed. By log scaling the data we find the distribution is close to normal around the mean, apart from minor outliers. Thus 𝒯_*t*_ and 𝒟_*t*_ represent the logarithm of the computed and reported values respectively.

In Equation 7, the ln ℒ‘s are computed for both infected cases and deaths and are then added.The likelihood has 2 error parameters, corresponding to the scatter in the infections and death data. The parameter *σ* appearing in the intrinsic scatter term encodes both parameters *σ*_1_ and *σ*_2_, corresponding to scatters in detected infection and death reports on a log scale. They appear in these two likelihood terms as defined in Equation 7. Note that the correlation in the data at different times can, in principle, be modelled through an error covariance matrix. We ignore such correlations here. In some cases reporting error leads to negative death numbers. Unless they are large (*>* 10) where we regularize the data by the absorption of outliers, we reject the data point.

#### 1.9.2 Priors

We use uniform priors on all the parameters. The ranges for each parameter are provided in Table 3. We ensure that the priors are broad enough to generate a two-tailed marginalized posterior distribution where the parameter can be constrained by the data.

**Table 3.**
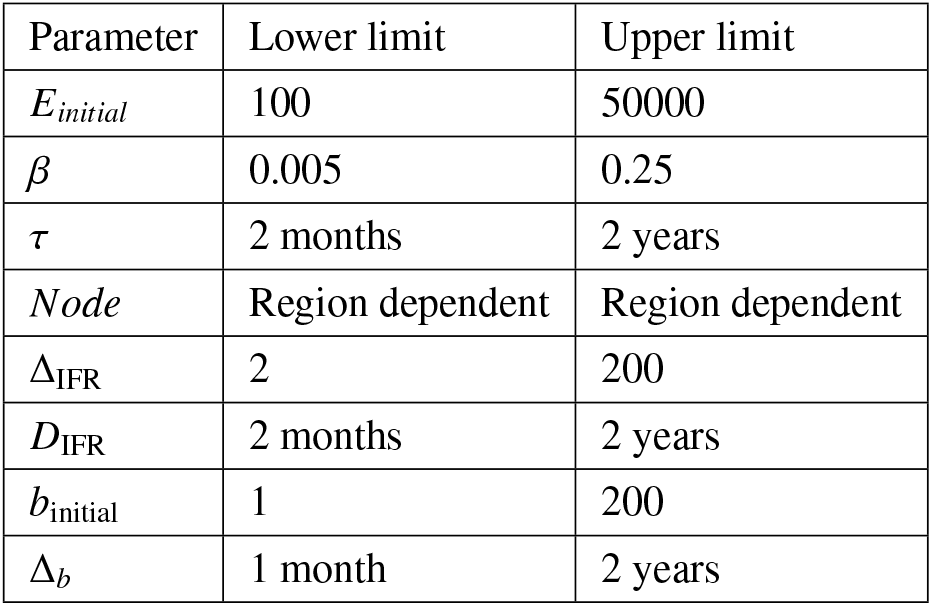
Priors on model parameters used in our analysis

Only for the period between nodes, the priors depend upon the zone that is being studied. With a visual inspection of the data, we can determine the possible times of transition. Different zones have local peaks at different time of the pandemic owing to the reasons described in subsection 1.8. To capture these effects, the priors on the nodes remain region dependent. However we impose wide priors around those transition times to remove possible bias. If the entire timeline is divided into *N* windows, we will have *N* − 1 nodes. The lower limit on the *i* + 1’th *Node* is the day following the upper limit of *i*’th *Node*.

#### 1.9.3 Parameter estimation and post processing

The posterior probability *p*(*θ* | *D, M*) for the parameter set *θ* of a model *M* given data D is given by Bayes Theorem,

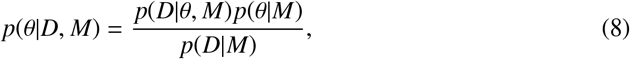

where *p*(*D* | *θ, M*) is the likelihood and *p*(*θ* | *M*) the prior probability distribution. *p*(*D* | *M*) is the Bayesian evidence or the marginal likelihood [91]. We use a nested sampling method for parameter estimation using `PolyChord` [94]. We consider uniform priors and the likelihood described in Equation 7 and use CosmoChord (an extension of CosmoMC that includes `PolyChord`) as our generic sampler [95]. A brief description of nested sampling with `PolyChord` is given in SI: Section 5.

While `PolyChord` computes the marginal likelihood, it also useful for parameter estimation using samples. We remove 30% of the total samples generated by the nested sampling procedure that represent the initial burn-in phase. We use getdist to obtain the marginalized posterior probabilities [96]. We use an adaptive parametrization as described in subsection 1.8. Marginal likelihoods are compared to determine the optimal numbers of windows required to describe the data. We show marginalized posteriors in certain cases in the main article while the others are presented in supplementary material.

### 1.10 Data

We use cumulative infected and death data downloaded from the COVID19India website https://www.covid19india.org/. We manually curated the time series, but only wherever required to deal with major outliers arising from reported corrections to the record. (For example Mumbai reported 917 deaths on June 16th 2020, as opposed to the average count of 78.3 for previous week.) These outliers were smoothed by replacing their values by the local average and redistributing the excess count over 20 - 60 previous days depending upon the value. Typically, for each time series there are only a handful of such outliers (*<* 5), if at all, and are removed manually. For our simulations, we need the population data for each zone of interest (district, state or country), and mention each of these sources in the appropriate sections.For each such zone, we also need the population fraction in each age group which we obtain from the 2011 census data [97].

### 1.11 Estimating mortality under-counting

Reported infections are expected to be substantially lower than the actual numbers of infected. While death numbers should nominally be better indicators of the state of the pandemic, most deaths in India happen at home and a medically certified cause of death may not be available, given low levels of MCCD coverage. Moreover, death registration is not uniform across the country. Estimates of overall under-counting of covid deaths vary, but reasonable estimates suggest a factor of 1.5 - 5 across India during the first wave [50].

Under-counting for cases and deaths cannot be estimated independently in our model, since one of these can be subsumed into a definition of the IFR. Benchmarking our results to those from serological surveys (serosurveys) provides a way of estimating this undercounting, since such surveys provide an estimate of the fraction of the population which has sustained a prior infection by the time of the test. Requiring that the quantum of deaths be consistent with the postulated IFR then allows us to estimate the undercounting of deaths at the time of the survey.

We analyse data for Karnataka - a typical Indian state with both rural and urban districts. (Approximately 62% of Karnataka’s population lives in rural areas, comparable to a figure of 65-70% for India as a whole.) We choose Karnataka for our analysis so that we can compare our numbers to district-wise serosurvey results available from a detailed study conducted in early September of 2020 [98].

For this part of the analysis, we assume that IFR does not change over the course of the first wave; we refer to this as the *fixed IFR* analysis. We choose several fixed values of (age-averaged) IFR in the range 0.15% and 0.3%. We vary the extent of death undercounting by multiplying the observed number of deaths by a factor 𝒰 *>* 1. We then chose a baseline district where it is potentially safe to assume that death under-counting might be minimal and that, hence, 𝒰= 1. For this district, we expect the number of infections as estimated from serosurveys should coincide with those obtained from our calculations as well as be consistent with the numbers of deaths given our assumed IFR.

Once we standardize our choice of (age-averaged) IFR with this method for a baseline district, we assume that the same IFR holds true across the state. We can then use the serosurvey results to estimate the true number of deaths for each district [98]. If *D*_*reported*_ is the number of reported deaths, *I*_*sero*_ is the expected number of infected individuals based on serosurvey, we can in principle estimate the death under-counting factor 𝒰 using the relationship between these quantities: 𝒰 = max[*IFR* * *I*_*sero*_*/D*_*reported*_, 1]. This relation is an average estimation. However, as the pandemic is ongoing during the serosurvey, the infection and deaths differ by a lag (≈ 1-2 weeks) and thus, we cannot use this relation directly.

Instead, using time series of reported infections and deaths, we first estimate actual infections by the time of serosurvey (*X*(*Z*) for a district *Z*). If there is no death under-counting, the expected proportion of actual infections will be equal to the actual proportion of infected from serosurveys (*Y*(*Z*)). We estimate any offset from this expectation to estimate a death under-counting factor for each district *Z*: 𝒰 (*Z*) = max[*Y*(*Z*)*/X*(*Z*), 1]. All these estimates were performed by analysing time-series data that begins with an initial simulation date (two weeks prior to the available infected and death reports of each district) and goes on to early September 2020, when the Karnataka serosurvey was conducted. We simultaneously fit the daily time-series of infection and deaths, using the fixed values of the (age-averaged) IFR mentioned above. Uncertainties in the serosurvey results translate to uncertainties in our determination of 𝒰 (*D*). Apart from the estimation of death undercounting, the rest of our analysis is based on a time-varying IFR described in the previous sections.

## 2 Results

We present our results in the following sequence. We begin with an estimation of death under-counting in Karnataka (subsubsection 2.1.1), using a simultaneous fitting of reported infections and deaths to our INDSCI-SIM model, and then comparing with serosurvey findings, following the fixed IFR analysis protocol described in subsection 1.11. We then apply our model allowing for time-varying IFR (and other parameters, as described in subsection 1.1 to 1.10), to data of each district of Karnataka (subsubsection 2.1.2).

This is followed by an analysis of data for a number of Indian cities where we assume no death undercounting (subsection 2.2). While this assumption can certainly be questioned, we do so on the grounds that the official numbers for deaths in the large Indian cities should be subject to smaller levels of undercounting than in rural India. We go on to model aggregate state-level data for Karnataka (subsection 2.3), assuming that our undercounting estimates for each district can be averaged to obtain a single number for the undercounting at the state level. Finally we apply the model to data for India as a whole, with death undercounting taken over from our estimates for Karnataka (subsection 2.4). In each of these cases, we estimate, among other parameters, the actual number of infected and IFR over the course of first wave of the pandemic.

### 2.1 Karnataka districts: Estimating death under-counting

#### 2.1.1 Estimates of death under-counting with fixed IFR

We use the serosurvey results published in Ref. [99] for all districts of Karnataka. We first scale all age-dependent IFR’s with a constant factor to ensure age-averaged IFRs of 0.3%, 0.2% and 0.15% – these have been argued to be in the right range for LMICs [80]. We assume further that either (a) there is no death under-counting or, (b) that deaths have been under-counted by a factor of 2. As can be seen in Table 4 assuming that deaths have been under-counted by a factor of two doubles our estimate of the actual infected in the same IFR category, as expected. On the other hand a decrease in the IFR results in an increase in our estimate of the actual numbers of infected. We stress that, for the purposed of this specific analysis, the IFR is assumed to be constant over time (no change in *δ*^*i*^) and that we only analyze the data till September 15, 2020 for all districts in Karnataka for consistency with the Karnataka serosurvey. Population data for Karnataka districts is sourced from the projections for the period 2020-2021, as provided by the report issued by Directorate of Economics and Statistics, Bangalore [100].

**Table 4.** Estimation of death under-counting by comparing seroprevalence. The first column lists each district of Karnataka (as on Sept 2020). The second column provides the seropositivity estimate as obtained in a serosurvey across the first two weeks of September 2020. The third column provides our estimate of the potential death undercounting in that district, 𝒰 (*D*), represented by the ratio of seropositivity to the actual number of reported infected. Since death overcounting is not possible, the minimum value of 𝒰 (*D*) can only be 1. The uncertainty on this number is obtained by propagating the 95% uncertainties from the serosurvey reports to our prediction of actual infection. In cases the lower bound on 𝒰 (*D*) goes below 1, we present the mean value and the range from 1 as an upper bound in parentheses. In a few cases we just present 1 as an upper bound, representing those cases where even the upper limit is less than or equal to 1.

|  |  | IFR=0.3%<br>No death multiplier |  | IFR=0.3%<br>Death multiplier=2 |  | IFR=0.2%<br>No death multiplier |  | IFR=0.2%<br>Death multiplier=2 |  | IFR=0.15%<br>No death multiplier |  |
| --- | --- | --- | --- | --- | --- | --- | --- | --- | --- | --- | --- |
| District | Observed<br>sero<br>prevalence (%) | Predicted<br>actual<br>infection | $\mathcal{U}(D)$ | Predicted<br>actual<br>infection | $\mathcal{U}(D)$ | Predicted<br>actual<br>infection | $\mathcal{U}(D)$ | Predicted<br>actual<br>infection | $\mathcal{U}(D)$ | Predicted<br>actual<br>infection | $\mathcal{U}(D)$ |
| Bengaluru Urban | 29.8 ± 3.3 | 19 ± 6.5 | 1.6 [1 - 2.3] | 34.3 ± 10.5 | 1 [1-1.3] | 26.9 ± 8.1 | 1 [1-1.6] | 52.1 ± 16.4 | 1 | 33.2 ± 8.3 | 1 [1-1.2] |
| Mysuru | 27.2 ± 8.8 | 13.9 ± 5.9 | 2 [1-3.5] | 25.7 ± 8.5 | 1.1 [1-1.8] | 19.7 ± 7.4 | 1.4 [1-2.4] | 37.2 ± 11 | 1 [1-1.2] | 25.7 ± 8.9 | 1.1 [1-1.8] |
| Ballari | 43.1 ± 9.5 | 13.6 ± 6.7 | 3.2 [1-5.5] | 25 ± 11.1 | 1.7 [1-2.8] | 19.5 ± 8.5 | 2.2 [1-3.7] | 34.9 ± 11.8 | 1.2 [1-1.9] | 25.4 ± 11.1 | 1.7 [1-2.8] |
| Dakshina Kannada | 27 ± 8.5 | 12.4 ± 4.9 | 2 [1-3.7] | 23 ± 7.7 | 1.2 [1-2] | 17.5 ± 5.9 | 1.5 [1-2.5] | 33.3 ± 10.6 | 1 [1-1.3] | 22.6 ± 7.4 | 1.2 [1-2] |
| Hassan | 30.7 ± 9.4 | 10.6 ± 6.7 | 2.9 [1-5.6] | 19.8 ± 11.4 | 1.6 [1-3] | 15 ± 9.1 | 2 [1-3.9] | 26 ± 12.6 | 1.2 [1-2.1] | 20.1 ± 11.6 | 1.5 [1-2.8] |
| Belagavi | 30.1 ± 8.8 | 4.2 ± 2.2 | 7.2 ± 5.8 | 8.3 ± 4.1 | 3.6 [1-6.5] | 6.2 ± 3.1 | 4.9 ± 3.8 | 11.4 ± 5.2 | 2.6 [1-4.6] | 7.9 ± 3.7 | 3.8 [1-6.7] |
| Tumakuru | 29.4 ± 9.5 | 5 ± 2.4 | 5.9 ± 4.7 | 9.4 ± 3.9 | 3.1 [1-5.4] | 7.4 ± 3.4 | 4 [1-7.1] | 14 ± 5.4 | 2.1 [1-3.6] | 9.2 ± 3.7 | 3.2 [1-5.5] |
| Udupi | 36.4 ± 9 | 7.3 ± 3.3 | 5 ± 3.5 | 15.5 ± 7 | 2.3 [1-3.9] | 11.3 ± 4.7 | 3.2 ± 2.1 | 22.2 ± 9.1 | 1.6 [1-2.7] | 13.9 ± 5.8 | 2.6 [1-4.3] |
| Kalaburagi | 29.8 ± 8.6 | 7.9 ± 4 | 3.8 [1-6.8] | 14.7 ± 6.9 | 2 [1-3.5] | 10.7 ± 4.6 | 2.8 [1-4.8] | 22.5 ± 9 | 1.3 [1-2.2] | 14.7 ± 6.6 | 2 [1-3.5] |
| Dharwad | 8.7 ± 6 | 18.9 ± 8.6 | 1 | 35 ± 11.8 | 1 | 26.5 ± 9.4 | 1 | 47.3 ± 12.2 | 1 | 36.6 ± 12.7 | 1 |
| Shivamogga | 21.4 ± 8.3 | 12.1 ± 5.4 | 1.8 [1-3.3] | 21.8 ± 9.2 | 1 [1-1.8] | 16.9 ± 7.5 | 1.3 [1-2.4] | 31.1 ± 12.5 | 1 [1-1.2] | 22.5 ± 9.3 | 1 [1-1.8] |
| Davanagere | 40.6 ± 9.7 | 16.6 ± 11.7 | 2.4 [1-4.7] | 28.7 ± 16.4 | 1.4 [1-2.5] | 22.7 ± 14.7 | 1.8 [1-3.4] | 36.8 ± 17.6 | 1.1 [1-1.9] | 29.5 ± 17.1 | 1.4 [1-2.5] |
| Mandya | 25.3 ± 8.6 | 5.3 ± 3.6 | 4.8 [1-9.7] | 9 ± 5 | 2.8 [1-5.3] | 7.9 ± 4.8 | 3.2 [1-6.2] | 13.6 ± 7.3 | 1.9 [1-3.5] | 9.1 ± 5.4 | 2.8 [1-5.4] |
| Bengaluru Rural | 28.7 ± 8.9 | 13.9 ± 9.7 | 2.1 [1-4.2] | 20 ± 11.7 | 1.4 [1-2.7] | 16.8 ± 10.9 | 1.7 [1-3.3] | 25.5 ± 12.5 | 1.1 [1-2] | 19.4 ± 11 | 1.5 [1-2.8] |
| Chitradurga | 25.9 ± 8.9 | 13 ± 9 | 2 [1-4.1] | 16.3 ± 8.4 | 1.6 [1-3] | 14.6 ± 8.4 | 1.8 [1-3.4] | 19.9 ± 8.2 | 1.3 [1-2.3] | 16.1 ± 8.3 | 1.6 [1-3] |
| Vijayapura | 35.4 ± 9.7 | 4.3 ± 1.6 | 8.2 ± 5.3 | 8.4 ± 2.9 | 4.2 ± 2.6 | 6.3 ± 2.1 | 5.6 ± 3.4 | 11.9 ± 3.6 | 3 ± 1.7 | 8.2 ± 2.6 | 4.3 ± 2.6 |
| Uttara Kannada | 16.3 ± 7.5 | 6.6 ± 3.8 | 2.5 [1-5.1] | 12.2 ± 6.5 | 1.3 [1-2.6] | 8.8 ± 4.5 | 1.9 [1-3.7] | 16.1 ± 7.2 | 1 [1-1.9] | 11.2 ± 5.2 | 1.5 [1-2.8] |
| Raichur | 34.1 ± 9.3 | 6.6 ± 2.3 | 5.2 ± 3.2 | 12.3 ± 3.6 | 2.8 ± 1.6 | 9.3 ± 2.7 | 3.7 ± 2.1 | 18 ± 4.6 | 1.9 [1-2.9] | 12.1 ± 3.2 | 2.8 ± 1.5 |
| Chikkamagaluru | 31.8 ± 9 | 9.8 ± 5.7 | 3.2 [1-6] | 16.1 ± 7.2 | 2 [1-3.4] | 13.4 ± 6.6 | 2.4 [1-4.2] | 23.7 ± 10 | 1.3 [1-2.2] | 18 ± 9.4 | 1.8 [1-3.2] |
| Koppal | 22.3 ± 7.9 | 15.9 ± 8.3 | 1.4 [1-2.6] | 29.1 ± 12.6 | 1 [1-1.4] | 23.5 ± 11.1 | 1 [1-1.7] | 38 ± 14.7 | 1 | 28.9 ± 12.5 | 1 [1-1.4] |
| Chikkaballapura | 12.1 ± 7.6 | 7.3 ± 4.5 | 1.7 [1-3.8] | 11.6 ± 5.3 | 1 [1-2.1] | 9.5 ± 5 | 1.3 [1-2.8] | 16.5 ± 6.3 | 1 [1-1.4] | 12.3 ± 5.8 | 1 [1-2.1] |
| Bagalkote | 12 ± 7.1 | 10.1 ± 7.5 | 1.2 [1-2.8] | 14.4 ± 8.5 | 1 [1-1.8] | 13.9 ± 9.5 | 1 [1-2] | 22.3 ± 12.2 | 1 [1-1.1] | 15.1 ± 9 | 1 [1-1.7] |
| Gadag | 9 ± 8 | 8.7 ± 3.6 | 1 [1-2.3] | 16.6 ± 5.7 | 1 [1-1.2] | 12.8 ± 4.6 | 1 [1-1.6] | 24.4 ± 8.3 | 1 | 16.5 ± 5.4 | 1 [1-1.2] |
| Haveri | 28.6 ± 8.8 | 10.7 ± 6.2 | 2.7 [1-5.1] | 17.9 ± 8.6 | 1.6 [1-2.8] | 14.4 ± 7.2 | 2 [1-3.6] | 24 ± 9.8 | 1.2 [1-2.1] | 18 ± 8.2 | 1.6 [1-2.8] |
| Yadgir | 31.6 ± 8.9 | 10.5 ± 7.9 | 3 [1-6.1] | 14.7 ± 9.6 | 2.1 [1-4.1] | 12.4 ± 8.5 | 2.5 [1-5] | 18.7 ± 9.9 | 1.7 [1-3.1] | 15.1 ± 9.3 | 2.1 [1-4] |
| Kolar | 16.1 ± 7.4 | 8.5 ± 6.3 | 1.9 [1-4.2] | 12.2 ± 7.5 | 1.3 [1-2.7] | 10.8 ± 7.5 | 1.5 [1-3.2] | 15.3 ± 8.3 | 1.1 [1-2.2] | 12.3 ± 7.3 | 1.3 [1-2.7] |
| Bidar | 18.7 ± 7.6 | 8.3 ± 4.6 | 2.3 [1-4.5] | 14.3 ± 6 | 1.3 [1-2.4] | 11.6 ± 5.7 | 1.6 [1-3] | 19.3 ± 6.9 | 1 [1-1.7] | 14.6 ± 5.9 | 1.3 [1-2.6] |
| Ramanagara | 29.3 ± 9.2 | 7.5 ± 4.5 | 3.9 [1-7.5] | 13.1 ± 6.2 | 2.2 [1-4] | 10.7 ± 6.1 | 2.7 [1-5.1] | 18.2 ± 7.7 | 1.6 [1-2.8] | 13.1 ± 6 | 2.2 [1-3.9] |
| Chamarajanagara | 21.1 ± 8.4 | 8.9 ± 6.5 | 2.4 [1-5.1] | 12.1 ± 6.4 | 1.7 [1-3.3] | 10.1 ± 5.9 | 2.1 [1-4.2] | 16.5 ± 7.1 | 1.3 [1-2.4] | 13 ± 7.2 | 1.6 [1-3.1] |
| Kodagu | 20.5 ± 8.2 | 14.9 ± 10.6 | 1.4 [1-2.9] | 19.8 ± 10.7 | 1 [1-2] | 16.6 ± 10 | 1.2 [1-2.4] | 24.4 ± 10.3 | 1 [1-1.5] | 19.5 ± 10.8 | 1.1 [1-2.1] |

We choose Bengaluru Urban as our baseline model district, assuming that the recorded numbers of deaths are accurate. This choice is motivated by the observation that Bengaluru Urban is the largest district in Karnataka by population. It contains the state capital and is also thus likely under greater scrutiny than other, more remote, districts. Comparing our model results with those from the serosurveys for Bengaluru Urban we find that an IFR of 0.2% (highlighted column in tha table) yields numbers of those actually infected that are close to the measured seroprevalence (the undercounting factor 𝒰 ≃1.1). To the extent that the reported death numbers are accurate for Bengaluru Urban, this is then our best estimate for the IFR – but only up to the date of the serosurvey. We can now use this assumption to estimate levels of undercounting in other districts, 𝒰 (*D*). (We, however, expect that this IFR value will go down further as time goes on, reflecting improvement in patient treatments and a consequent mortality reduction as well as COVID deaths in older age-bands leading to a reduction in the population which carries the most mortality risk.)

Using these columns in Table 4, for *IFR* = 0.2%, we plot the factors 𝒰 (*D*) in Figure 3 as a choropleth in the left panel. Darker colors for districts represent a larger death undercounting. In the right panel we plot 𝒰 (*D*) for *IFR* = 0.3% analysis. As expected, the estimated death undercounting increases here. This would be consistent with an death undercounting in the Bengaluru Urban district by 60% i.e. (U(*D*) = 1.6).

**Fig 3.**
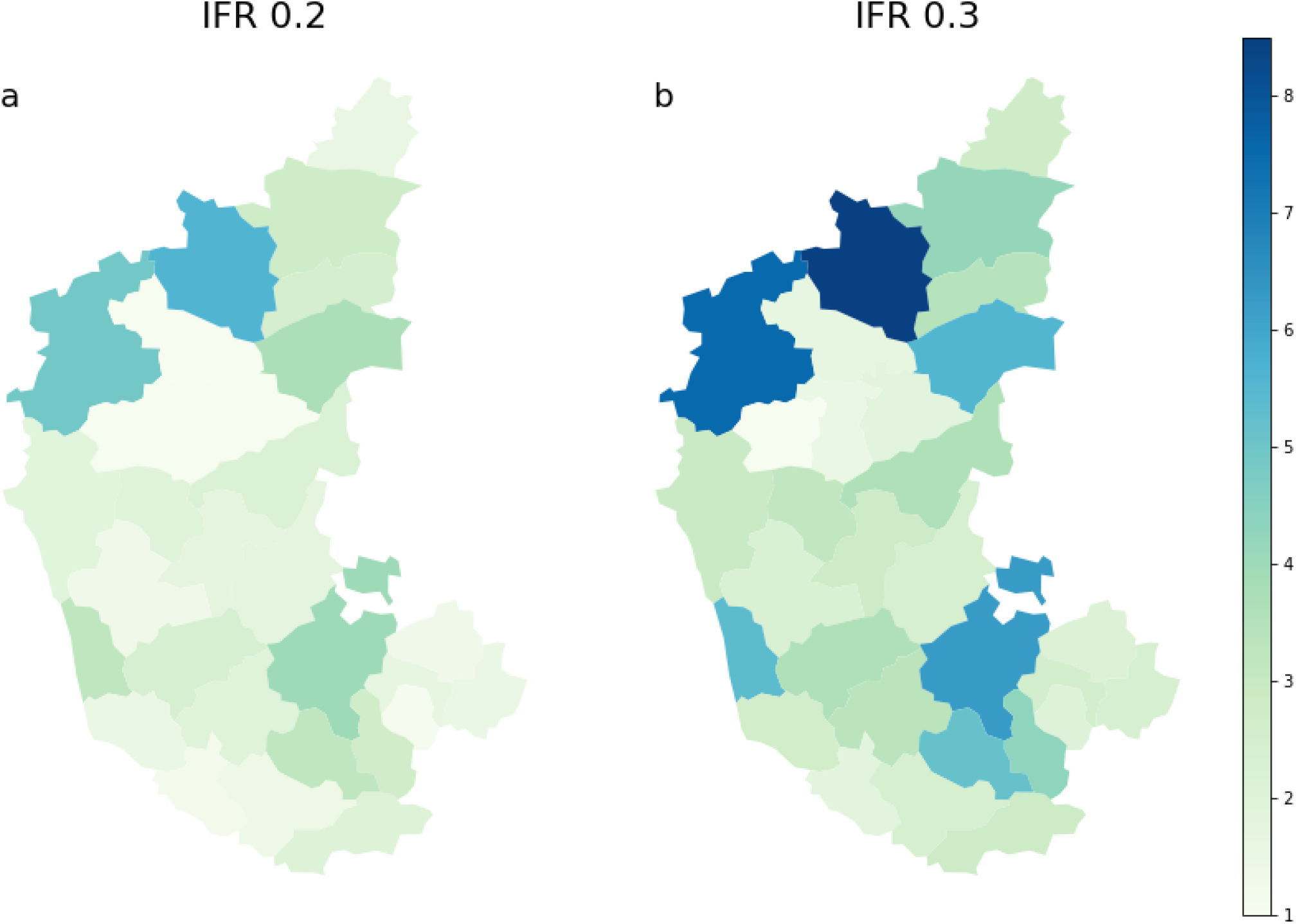
Ratio of seroprevalence reported in districts of Karnataka across the first two weeks of September, 2020 and our prediction of actual infected on the same date. This ratio indicates the death under-counting in different districts assuming uniform IFR in different districts. We plot the ratios here for IFR = 0.2 [a: left plot] and 0.3 [b: right plot] respectively. Note that these numbers are plotted from Table 4 for corresponding IFRs. For IFR = 0.3 the INDSCI-SIM prediction of actual infected is certainly lesser than the IFR = 0.2 case. The plot on the right shows larger ratios compared to the left.

#### 2.1.2 Estimates of cases by the end of first wave, with variable IFR

Having estimated 𝒰 (*D*), we redo our analysis for each of Karnataka’s districts, multiplying reported deaths by the multiplying factors tabulated in Table 4. We then run the program forward till February 15, 2021, but now also allow the IFR to vary according to the parametrization described earlier. Using this analysis we tabulate the actual infected percentages we predict by Dec 15, 2020 and Feb 15, 2021 in Table 5, together with 95% confidence intervals. We find that between 20% and 70% of the population in these districts has been infected by February 15, 2021. We find no substantial change in the actual infection between December 2020 and February 2021.

**Table 5.** Estimation of actual infection in Karnataka districts on December 15, 2020 and February 15, 2021. The uncertainties are quoted at 95% C.L. This table is obtained from an analysis that assumes a time varying IFR as explained in Methods.

| District | December 15, 2020 | February 15, 2021 |
| --- | --- | --- |
| Bengaluru Urban | 73.5 ± 10.5 | 74.5 ± 10.5 |
| Mysuru | 34.6 ± 10.9 | 36.2 ± 11.4 |
| Ballari | 46.1 ± 13.3 | 47.5 ± 13.4 |
| Dakshina Kannada | 38.0 ± 11.7 | 39.6 ± 12.4 |
| Hassan | 37.0 ± 12.4 | 38.0 ± 12.8 |
| Belagavi | 33.2 ± 8.3 | 34.1 ± 8.4 |
| Tumakuru | 45.9 ± 13.2 | 48.9 ± 14.5 |
| Udupi | 48.7 ± 20.0 | 50.3 ± 20.4 |
| Kalaburagi | 41.1 ± 14.7 | 42.0 ± 15.0 |
| Dharwad | 33.7 ± 13.1 | 35.0 ± 13.5 |
| Shivamogga | 25.3 ± 9.4 | 25.9 ± 9.7 |
| Davanagere | 50.4 ± 22.4 | 51.3 ± 22.9 |
| Mandya | 38.3 ± 19.0 | 39.1 ± 19.5 |
| Bengaluru Rural | 50.4 ± 20.7 | 52.4 ± 21.3 |
| Chitradurga | 36.7 ± 22.9 | 38.7 ± 23.8 |
| Vijayapura | 43.7 ± 9.9 | 44.7 ± 10.1 |
| Uttara Kannada | 34.7 ± 15.2 | 35.8 ± 15.8 |
| Raichur | 43.3 ± 13.0 | 44.4 ± 13.0 |
| Chikkamagaluru | 67.8 ± 18.6 | 69.6 ± 19.0 |
| Koppal | 43.7 ± 25.8 | 43.6 ± 26.1 |
| Chikkaballapura | 28.8 ± 16.4 | 30.7 ± 17.6 |
| Bagalkote | 33.3 ± 27.1 | 33.6 ± 27.3 |
| Gadag | 37.6 ± 27.9 | 38.2 ± 28.3 |
| Haveri | 30.0 ± 10.8 | 31.1 ± 11.0 |
| Yadgir | 34.9 ± 19.2 | 35.5 ± 19.6 |
| Kolar | 21.3 ± 8.5 | 22.0 ± 9.0 |
| Bidar | 24.5 ± 9.3 | 25.4 ± 9.4 |
| Ramanagara | 40.3 ± 15.4 | 40.8 ± 15.6 |
| Chamarajanagara | 38.2 ± 17.6 | 39.6 ± 18.2 |
| Kodagu | 45.5 ± 22.8 | 49.2 ± 24.3 |

The relationship between actual numbers of prior infections and results from serosurveys is complicated by the observation that antibodies measured in such surveys have been observed to decay in concentration over a period of several months. However, given the fact that the peak in cases in Karnataka occurred over September-October 2020, and that the serosurvey was performed during this period, it is reasonable to assume that this will not impact our interpretation of the serosurvey observations [99].

### 2.2 Infection curves for select Indian cities

In this sub-section, we provide an analysis of the progress of COVID-19 across the period Jan 2020 - February 15, 2021, for a few select Indian cities: Bengaluru, Chennai, Delhi, Pune and Mumbai. For each of these cities, the data correspond to the urban district agglomeration within which the city is embedded. We assume that deaths have been counted largely accurately for these cities, and thus that *U*(*D*) defined above is 1. We will return to the implications of this assumption later.

All these cities show more than a single peak in reported cases. The origins of these peaks are complex. Together, they represent a combination of relaxations of the lockdown and of physical distancing, festivals and political activity at various times, the ebb and flow of migration from nearby districts, as well as day-to-day variations in testing. These effects are captured in our multiple window model with adaptive selection, since infectivity parameters can now vary across different windows and the bias factor between actual and reported cases can itself evolve.

For each city, we present a set of 6 plots in three panels. In the top panel we present the fit to the daily reported cases (left - (a)) and to the daily reported deaths (right - (b)). The actual predicted infections and the bias are plotted in the middle left (c) and right (d) plots respectively. Note that the bias factor, once multiplied with the reported infections, provides the estimate of actual infection. The change in the IFR (left - (e)) and the effective reproduction number *R*(*t*) (right - (f)) estimates are plotted in bottom panel. For *R*(*t*) we also plot an independent estimate [84] as a reference.

#### 2.2.1 Bengaluru

Our analysis for Bengaluru Urban is presented in Figure 4; posterior distributions and parameter constraints are provided in SI: Section 3, Figure 2 and Table 1). For the calculations, the population data for Bangalore is sourced from the projections for the period 2020-2021, as provided by the report issued by Directorate of Economics and Statistics, Bangalore [100]. Our approach yields a double-peak structure in the curve of daily infections, reflecting a temporary flattening of the curve around September 2020 followed by an increase that resulted in a peak around October. Between mid-September and mid-October, we observe a plateau in deaths. Our results can be compared to those from serosurveys, as quoted in Refs. [101–103]. In September of 2020, these calculations yield a seroprevalence in Bengaluru Urban of about 30%, consistent with serosurvey results.

**Fig 4.**
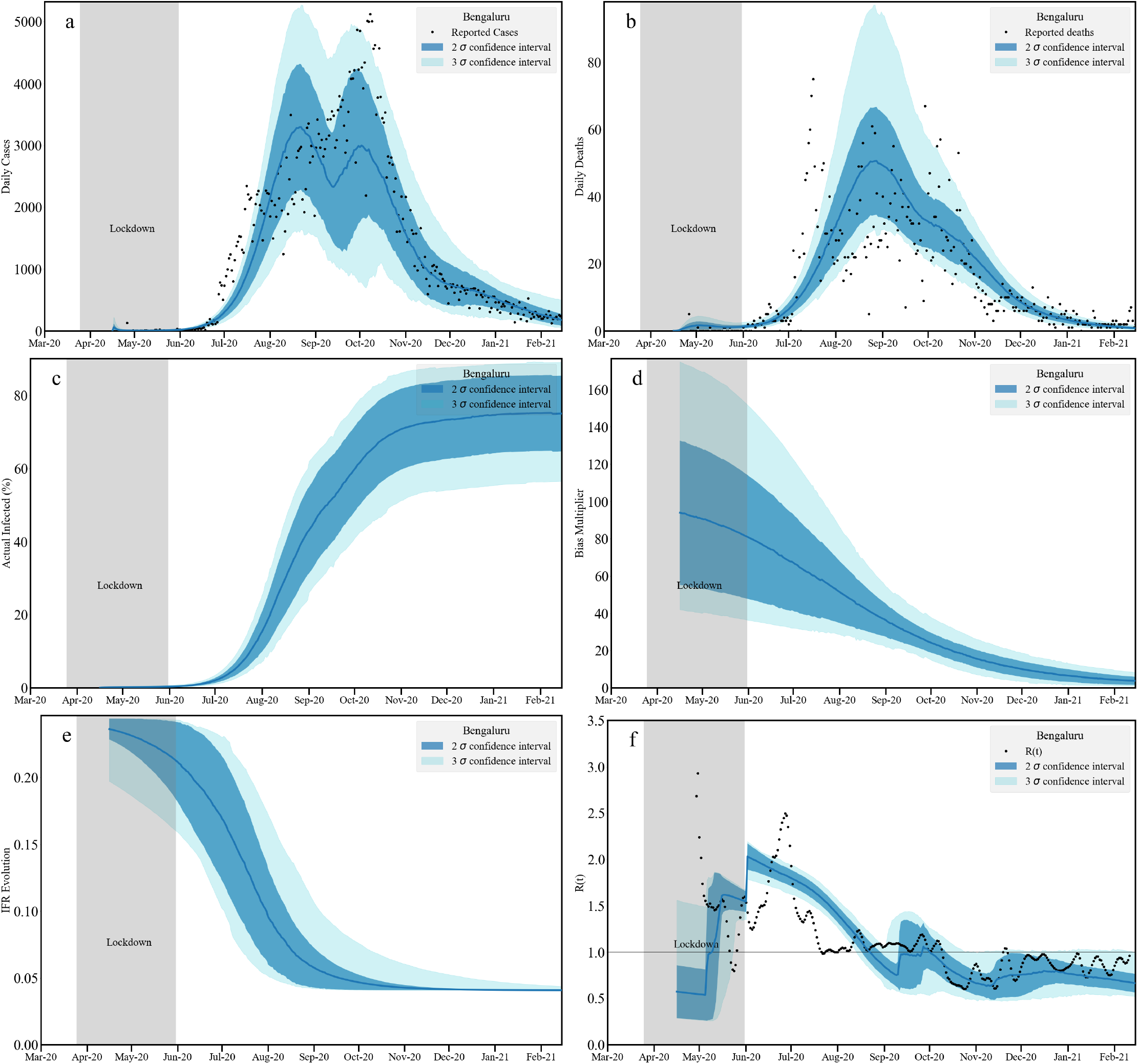
Timeseries analysis for Bengaluru Urban. We plot the fit to the daily infected cases [a: top left] and daily reported deaths [b: top right] assuming no death undercounting. The middle panel contains the cumulative actual infected cases [c: left] and the bias multiplicative factor [d: right] obtained as a ratio between actual and reported infections. The left plot [e] at the bottom panel contains the evolution of the age averaged IFR. The bottom right plot [f] contains our estimation of *R*(*t*) and an independent [84] measurement. Note that the bands correspond to 2*σ* and 3*σ* confidence levels.

We note that the model indicates that a substantial fraction of the population in Bengaluru had already been infected by Feb 15. The effective reproductive ratio *R*(*t*) decreases abruptly during the lockdown, while displaying an equally sharp increase post lockdown. Our estimate for those infected in Bengaluru city by Feb 15, 2021 lies between 64% and 85% at the 95% confidence level. The bias multiplier narrows quite rapidly, reading a figure of about 10 by mid-February 2010; at the time of the Bengaluru serosurvey, this factor was approximately 30. Our estimate for the IFR at the end of the first wave indicates a value in the vicinity of 0.05%, roughly consistent with similar estimates for the first wave based on serosurveys. An increase in the IFR, say to 0.08 or 0.1, can also be achieved if we assume that deaths have been under-counted by a suitable factor, while leaving our estimates for total infections the same. The cumulative infections and deaths are plotted together with the data in SI Section 3, Figure 8.

#### 2.2.2 Chennai

Our results for Chennai are plotted in Figure 5, with posterior distributions and parameter constraints provided in the SI: Section 3, Figure 3 and Table 2). For these calculations, we source population data for Chennai from Wikipedia, which takes into account the expansion of the district limits in 2018 [104]. Apart from the national lockdown of 68 days, we note that Chennai had imposed a shorter, local lockdown during the period June 19 - July 5, 2020. Both lockdown periods are marked in grey. While across the first lockdown the numbers of infected increased steadily, the numbers began to show a decline following the second lockdown. We see minor peaks around mid-August and October of a lesser height when compared to the first peak. This decrease is also corroborated by the estimates for the *R*(*t*). Serosurvey results for Chennai have been reported in Ref. [105].

**Fig 5.**
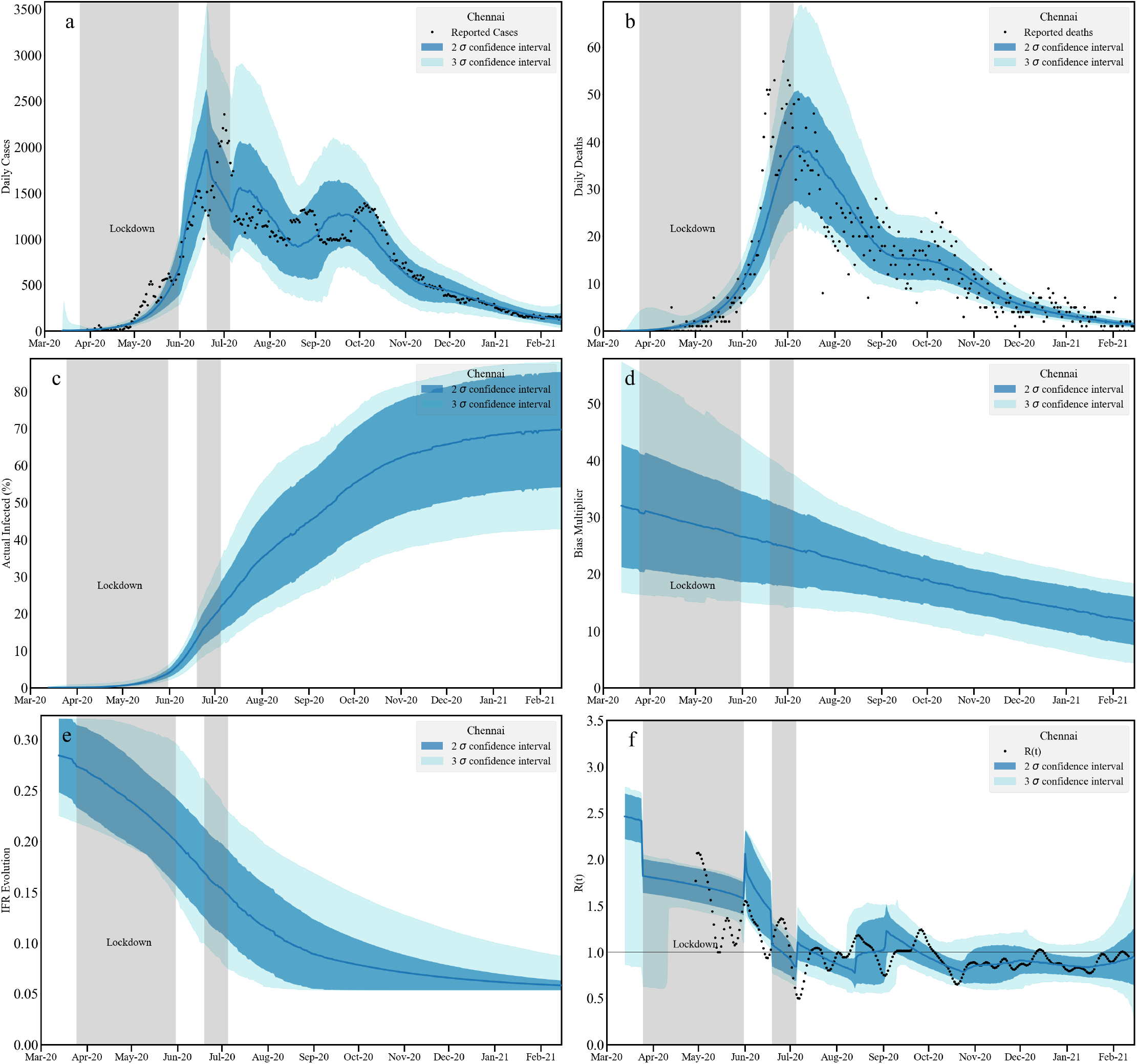
Timeseries analysis for Chennai. We plot the fit to the daily infected cases [a: top left] and daily reported deaths [b: top right] assuming no death undercounting. Middle panel contains the cumulative actual infected cases [c: left] and the bias multiplicative factor [d: right] obtained as a ratio between actual and reported infections. The left plot [e] at the bottom panel contains the evolution of age averaged IFR. The bottom right plot [f] contains our estimation of *R*(*t*) and an independent [84] measurement. Note that the bands correspond to 2*σ* and 3*σ* confidence levels.

There are clearly issues with data here since around July 2020, the peak in deaths appears to precede the infection peak, although the data is quite noisy. After that, a steady decrease in the death numbers is seen. While between mid-July to end-October the infection numbers change only marginally, the numbers of deaths reduce largely monotonically. This suggests an effective decrease in IFR, perhaps related to improvements in patient management. The bias multiplier also show a decrease. However the rate of the decline is much slower compared to Bengaluru Urban. The plot indicates a 30-fold undercounting of cases initially. This is reduced to around 15 by the middle of February 2020. A low test positivity since the onset of infection is supported by this plot and attributed to a large scale testing program. The actual infection plot suggests that nearly 54-85% (at 95% C.L.) of the Chennai population has been infected by mid-February.

We can compare these predictions to data from Ref. [46] for the state of Tamil Nadu as a whole in December 2020, which found that seroprevalence in urban areas (36.9%) was higher than in rural areas (26.9%). They found that 22.6 million persons were infected by the end of November, roughly 36 times the number of confirmed cases. For Chennai, the estimated seroprevalance by December was in the vicinity of 40%, within the 95% bound of our own results. The estimated seroprevalence implied an infection fatality rate of 0.052%, comparable to our estimates. Our estimates for cumulative infections and deaths are plotted together with the data in SI Section 3, Figure 9).

#### 2.2.3 Delhi

Figure 6 present our analysis with the Delhi data. The data for Delhi presents a challenge to modelling because of the presence of multiple peaks. The data requires 5 adaptive windows to fit. Within these 5 windows, the constrained infectivity parameter *β* show a oscillatory trend. This oscillatory pattern may possibly reflect patterns in the return of migrant workers to the national capital region. A number of different serosurvey results have been reported for Delhi, including in Ref. [106]. For our calculations, we use Delhi’s population data from the projections reported in the 2019 report published by the National Commission on Population [107].

**Fig 6.**
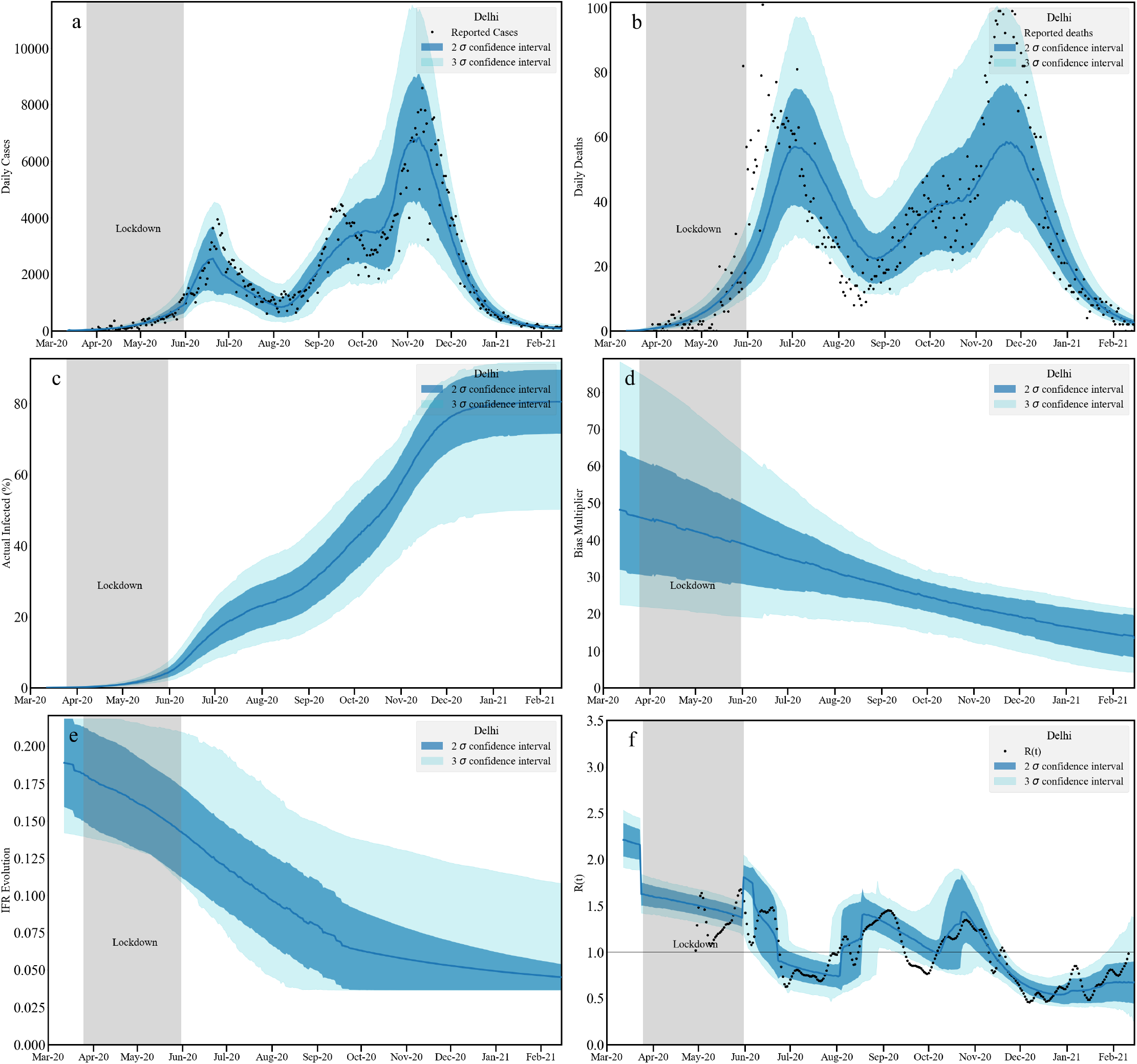
Timeseries analysis for Delhi. We plot the fit to the daily infected cases [a: top left] and daily reported deaths [b: top right] assuming no death undercounting. Middle panel contains the cumulative actual infected cases [c: left] and the bias multiplicative factor [d: right] obtained as a ratio between actual and reported infections. The left plot [e] at the bottom panel contains the evolution of age averaged IFR. The bottom right plot [f] contains our estimation of *R*(*t*) and an independent [84] measurement. Note that the bands correspond to 2*σ* and 3*σ* confidence levels.

With these windows, our model captures the trends in the data. Posterior distributions and parameter constraints are provided in the SI: Section 3, Figure 4 and Table 3. We find multi-peak posteriors in the *Node* parameters reflecting several possible solutions of the Delhi trajectory. The reported deaths here show outliers around June 2020 that occur before infection peaks towards the middle and end of June. Although the IFR decreases we find that it is still consistent with 0.1% IFR around February 2021. The bias multiplier settles down around 20, indicating that a point estimate of about 80% (71-90% at 95% C.L.) infected population in Delhi by January 2021, with a confidence of interval of ± 10%. Our limits at the 3*σ* confidence limit though indicate a range of 50-90% infected, given the skewed posterior distribution of bias. The estimated *R*(*t*) as expected show an oscillatory behavior, consistent with the independent estimates. We observe that *R*(*t*) fluctuates in a range *R*(*t*) ~ 1 − 1.5 before it reduces below 1 around December 2020.

Results from the fifth serosurvey in Delhi concluded that about 56% of the over 28,000 people whose blood samples were collected in January 2021 had developed antibodies against COVID-19. The first such survey done in the city in June-July had shown that 23.4% of people surveyed had developed antibodies against the virus. Similar surveys in August showed that 29.1% of people had antibodies to SARS-CoV-2 at that time. This became 25.1% in September and 25.5% in October. These results are approximately consistent with the results we present here, except for the January serosurvey, which lies outside our 95% confidence interval. However, due to the decay of antibodies, seroprevalence as derived from CLIA tests are expected to underestimate numbers of infected. We expect that accounting for this decay would yield closer agreement. Initial estimates of Delhi’s IFR in REf. [108] yield numbers consistent with a range (0.05-0.1%), consistent with our calculation here. However, other calculations yield a cumulative proportion of the population estimated infected of 48.7% (95% CrI 22.1% – 76.8%) by end-September 2020. These are noticeably larger than our own estimates of about 30% at that time. (The IFR assumed in that work was considerably larger, though, between 0.22% and 0.39%; this required that a relatively small number of deaths were actually recorded, about 28% of the actual number if the IFR was 0.21%.). Estimations for cumulative infections and deaths are plotted with data in SI (Ref. Section 3, Figure 10.

**Fig 7.**
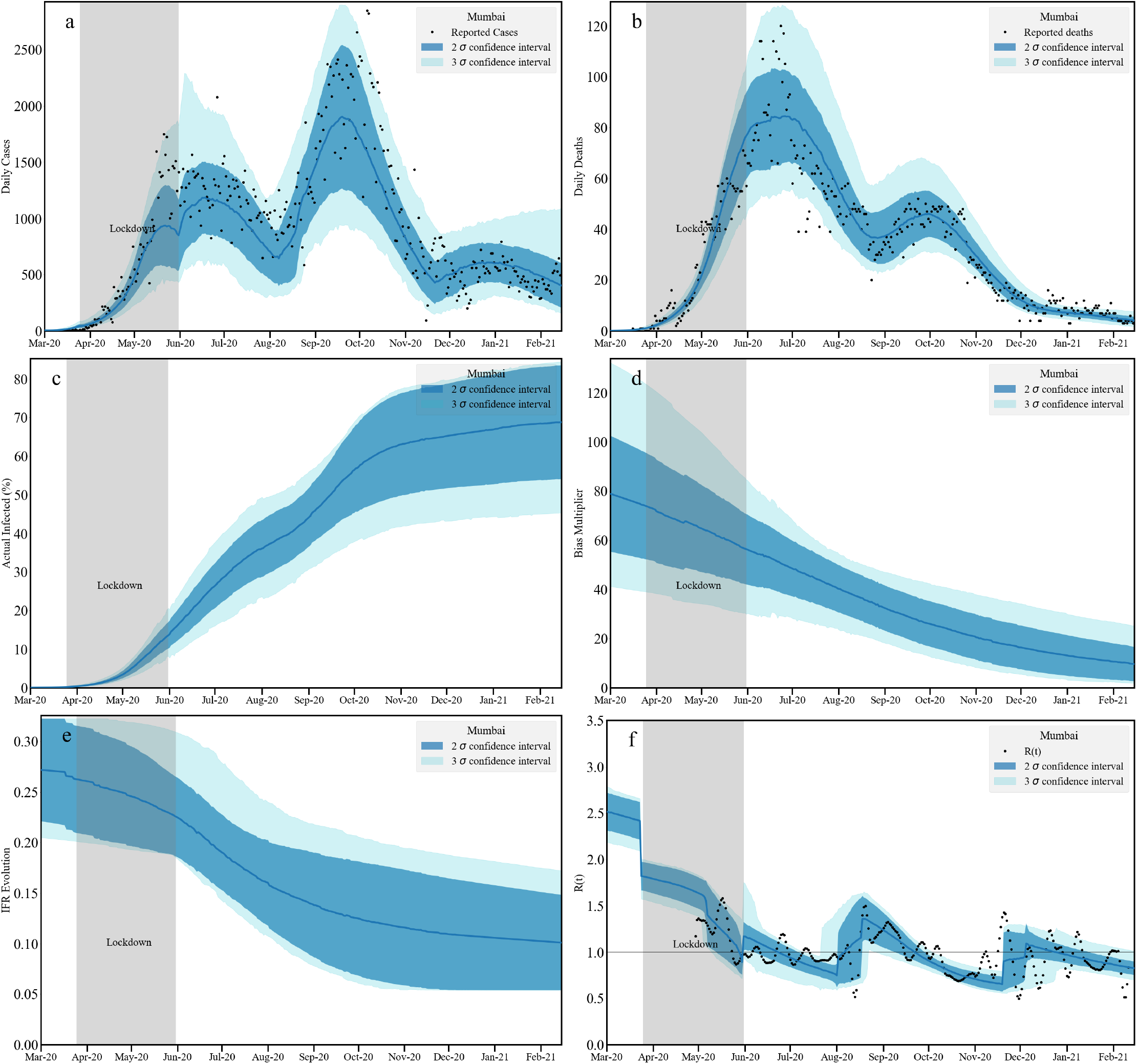
Timeseries analysis for Mumbai. We plot the fit to the daily infected cases [a: top left] and daily reported deaths [b: top right] assuming no death undercounting. Middle panel contains the cumulative actual infected cases [c: left] and the bias multiplicative factor [d: right] obtained as a ratio between actual and reported infections. The left plot [e] at the bottom panel contains the evolution of age averaged IFR. The bottom right plot [f] contains our estimation of *R*(*t*) and an independent [84] measurement. Note that the bands correspond to 2*σ* and 3*σ* confidence levels.

**Fig 8.**
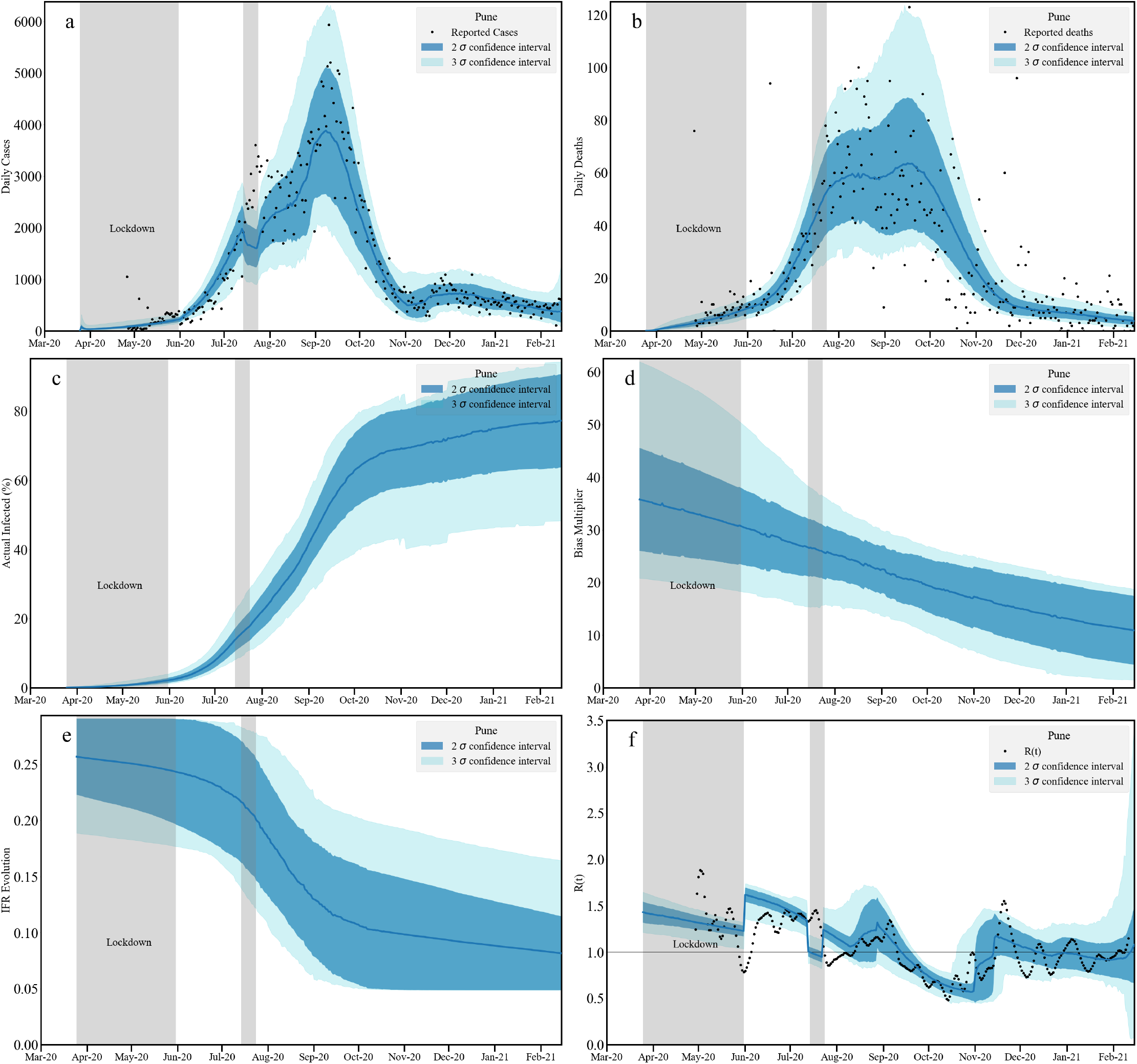
Timeseries analysis for Pune. We plot the fit to the daily infected cases [a: top left] and daily reported deaths [b: top right] assuming no death undercounting. Middle panel contains the cumulative actual infected cases [c: left] and the bias multiplicative factor [d: right] obtained as a ratio between actual and reported infections. The left plot [e] at the bottom panel contains the evolution of age averaged IFR. The bottom right plot [f] contains our estimation of *R*(*t*) and an independent [84] measurement. Note that the bands correspond to 2*σ* and 3*σ* confidence levels.

**Fig 9.**
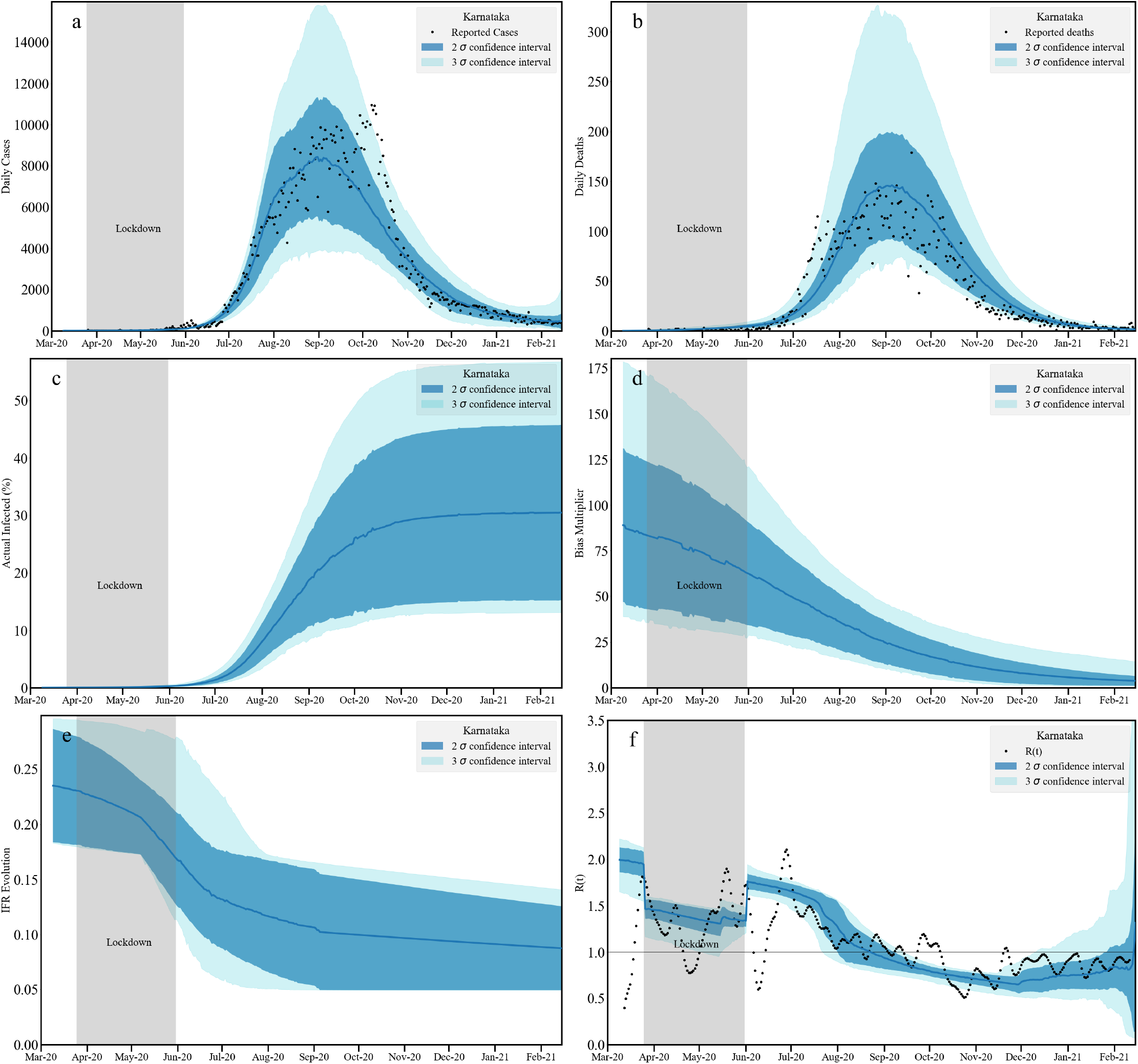
Timeseries analysis for Karnataka without death undercounting. We plot the fit to the daily infected cases [a: top left] and daily reported deaths [b: top right] assuming no death undercounting. Middle panel contains the cumulative actual infected cases [c: left] and the bias multiplicative factor [d: right] obtained as a ratio between actual and reported infections. The left plot [e] at the bottom panel contains the evolution of age averaged IFR. The bottom right plot [f] contains our estimation of *R*(*t*) and an independent [84] measurement. Note that the bands correspond to 2*σ* and 3*σ* confidence levels. We do not assume any death undercounting in this analysis.

**Fig 10.**
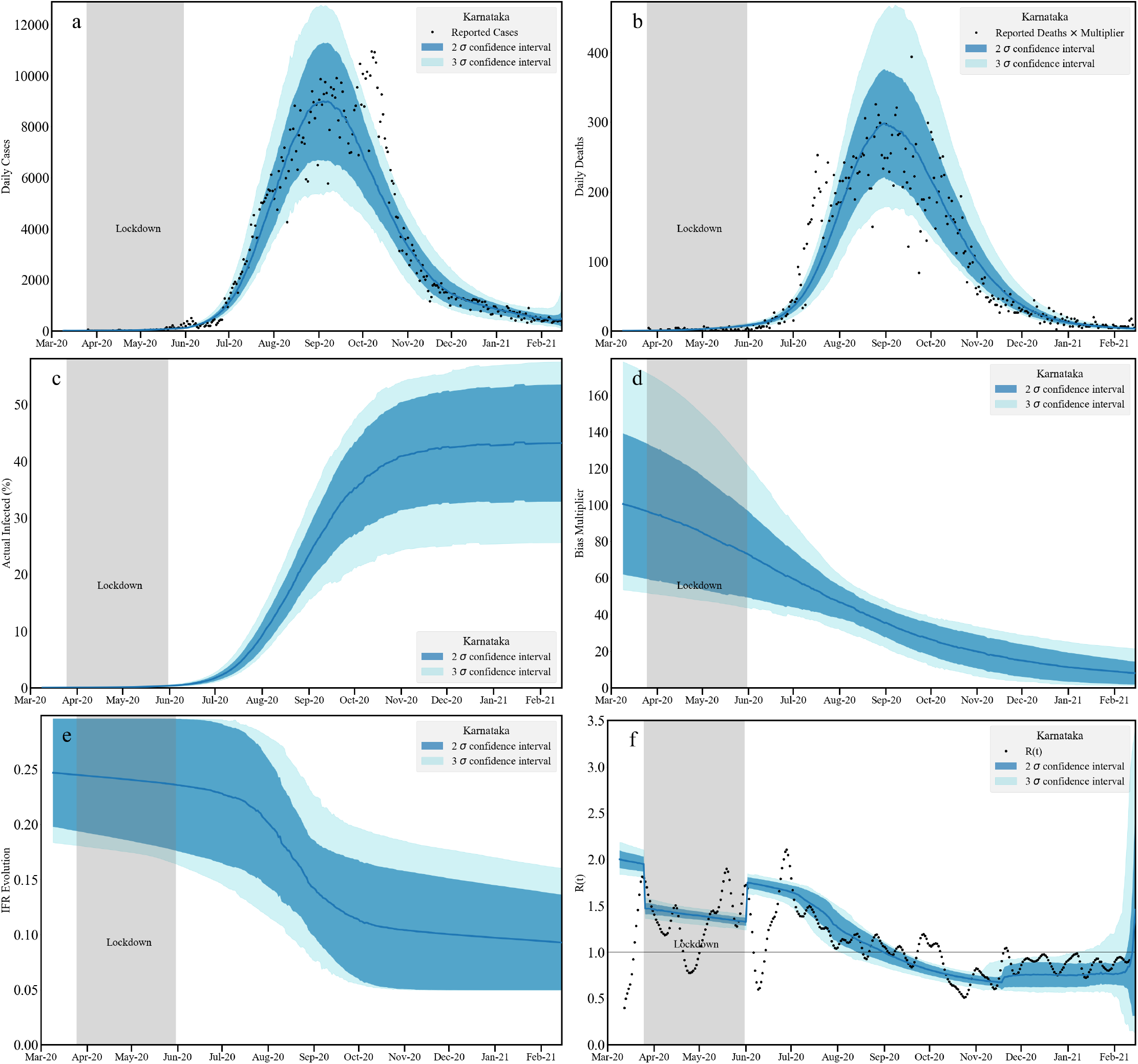
Timeseries analysis for Karnataka with death undercounting. We plot the fit to the daily infected cases [a: top left] and daily reported deaths [b: top right] taking into account estimated death undercounting. Middle panel contains the cumulative actual infected cases [c: left] and the bias multiplicative factor [d: right] obtained as a ratio between actual and reported infections. The left plot [e] at the bottom panel contains the evolution of age averaged IFR. The bottom right plot [f] contains our estimation of *R*(*t*) and an independent [84] measurement. Note that the bands correspond to 2*σ* and 3*σ* confidence levels. In this analysis the reported death numbers are multiplied with 2.2, the average death undercounting we obtained from Karnataka districts assuming Bengaluru Urban does not have any death undercounting.

#### 2.2.4 Mumbai

Results for the city of Mumbai are provided in Figure 7. The population of the district is taken from the projections for 2021 [109]. The posterior distributions and related parameter constraints are provided in the SI, Section 3, Figure 5 and Table 4. The data for Mumbai show two well resolved peaks around June-July 2020 and September-October 2020. Following the second peak, reported infections reduce and remain roughly constant from about December 2020. The reported deaths shows a peak and a plateau before reaching a second, smaller peak. While the second peak in the reported infection is higher than the first peak, the plateau in the reported death numbers around October 2020 is substantially lower than the July 2020 peak, indicating better treatment interventions. There are no systematic outliers in the death numbers, once we apply our smoothing to the data. The sharp decrease in death cases in contrast to the increase in reported infections could result from increase in testing (a lowering of the TPR) and/or a decrease in IFR.

While the bias multipler decreases from 100 to 15, the IFR does not decrease as rapidly. At 2*σ* the data is consistent at about an IFR of 0.15%, which is the largest when compared to the previously discussed cities. In Mumbai, a relatively smaller fraction of population (50%) appears to be infected within this calculation. However, fluctuations in the infected data *w*.*r*.*t*. the mean value translate to the uncertainties in the time-series estimation. Thus, the bands are wider compared to Bengaluru, Chennai and Delhi. At the 2*σ*-level we estimate that 54-84% of total Mumbai population may have been infected. *R*(*t*) shows a steady decrease within the lockdown and after that till mid-August 2020 when it starts to rise again temporarily indicating an upcoming second peak. Following the peak *R*(*t*) first reduces below 1 for a few months, then rises to fluctuate around 1 in early 2021 till February.

Mumbai serosurvey data showed that 54·1% of samples in slums and 16·1% of those in non-slums tested positive [110], in a study performed between June 29 and July 19, 2020. These are broadly consistent with our estimates. Our estimates for cumulative infections and deaths are plotted with data in the SI Section 3, Figure 11).

**Fig 11.**
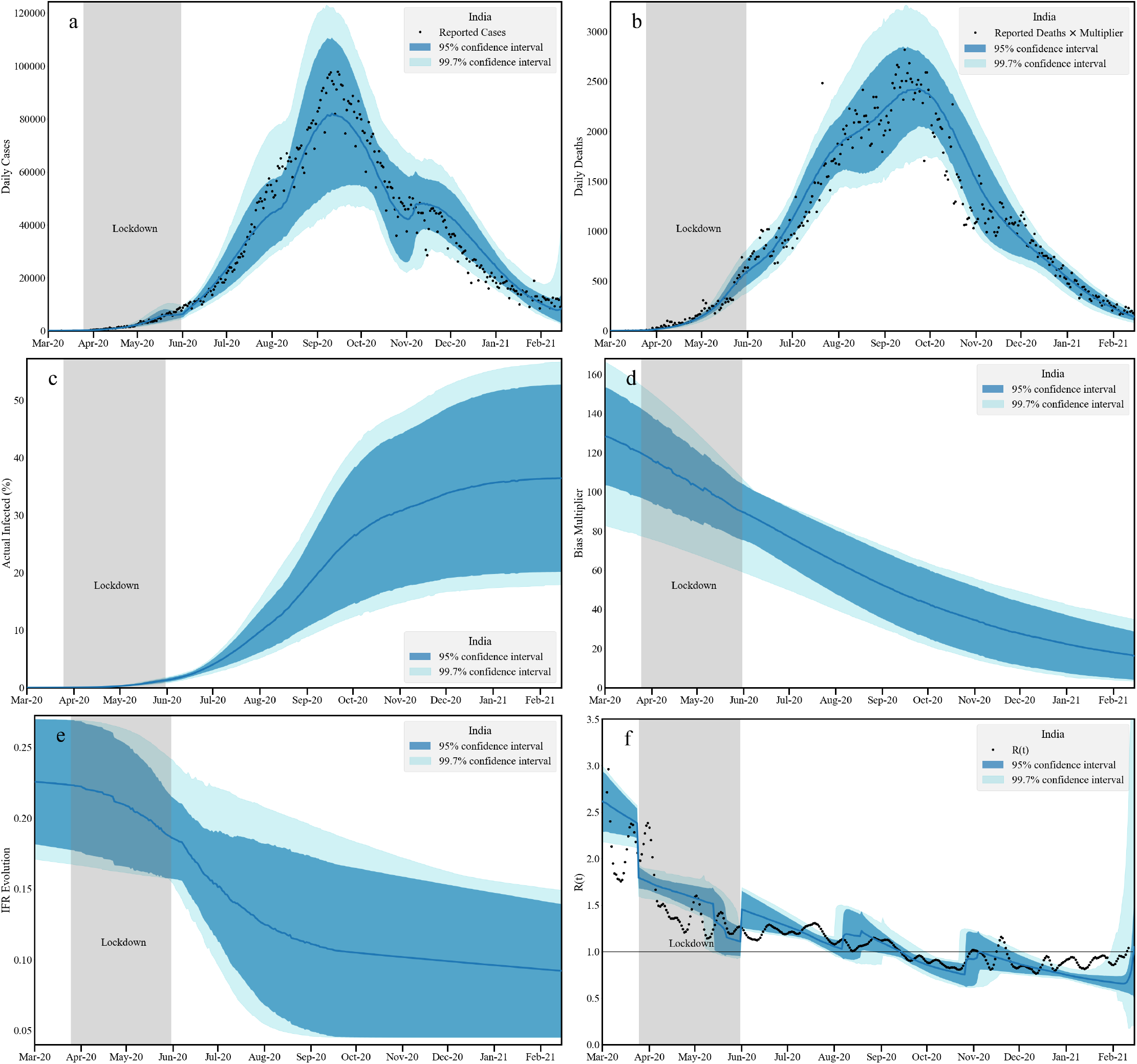
Timeseries analysis for Indian national data. We plot the fit to the daily infected cases [a: top left] and daily reported deaths [b: top right] assuming taking into account estimated death undercounting. Middle panel contains the cumulative actual infected cases [c: left] and the bias multiplicative factor [d: right] obtained as a ratio between actual and reported infections. The left plot [e] at the bottom panel contains the evolution of age averaged IFR. The bottom right plot [f] contains our estimation of *R*(*t*) and an independent [84] measurement. Note that the bands correspond to 2*σ* and 3*σ* confidence levels. In this analysis the reported death numbers are multiplied by 2.2 to take into account a measure of presumed undercounting

#### 2.2.5 Pune

In this section we present our final city analysis, that of Pune in Figure 8. The population of the district is taken as per projection for 2021 [109]. The posterior distributions and parameter constraints for this calculation are provided in SI: Section 3, Figure 6 and Table 5. As was the case with Chennai, Pune had imposed a local lockdown during 14-24 July 2020. There is substantial scatter in the reported death data, compared to the data for numbers of infected. This may arise from incomplete reporting initially. The infection data show a single prominent peak structure with a possible kink around mid-July 2020. The global peak comes around mid-September 2020, with infected numbers decreasing after that and becoming nearly constant around November. The death reports follow the infection in standard manner. However we do not find a substantial decrease in reported deaths *vis a vis*. infections. This is also reflected in the IFR plot where we see relatively minor changes from the initial IFR.

The bias multiplier changes from 35 to 15 indicating an improvement in testing. We also find a large infected fraction, of 64-90% of the population in Pune. Our estimated *R*(*t*) stays around 1.5 till September 2020 after which it comes down sharply to nearly 0.5 in two months. In December we see a rise in *R*(*t*) which remains nearly constant till the end of the first wave around mid-February. *R*(*t*) from December 2020 to February 2021 stays at nearly 1 which can also be observed in the flatness in reported cases and deaths.

A serosurvey in Pune, conducted between 20th July and 5th August 2020, found a seroprevalance of 51·3% (95%CI 39·9 to 62·4) [111]. The overall IFR was calculated to be 0.21. The inferred seroprevalence lies well above our own median prediction of about 35% by August 2020, although our IFR estimates at that time are closer to the value inferred from the serosurveys, at about 0.17. We note, however, that the substantial spread in the inferred IFRs across different localities even within the city make comparisons more difficult. Compartmental models are insensitive to such ultra-local variations; other, more refined individual-based models will be required to assess these effects. Estimations for cumulative infections and deaths are plotted with data in SI (Ref. Section 3, Figure 12).

**Fig 12.**
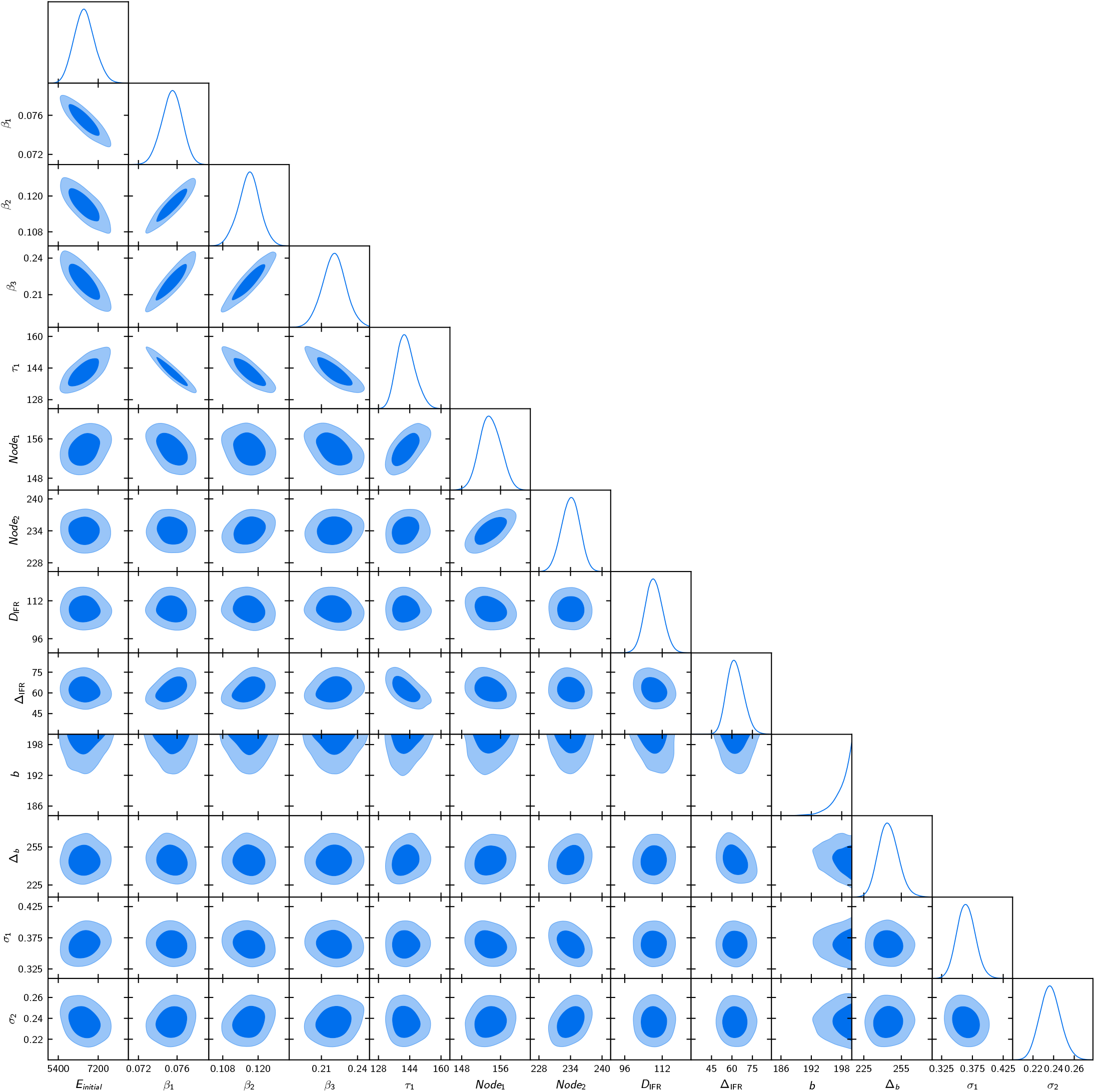
Marginalized posteriors of the parameters in the adaptive parametrization for India-level data.

### 2.3 Projections and data fitting for Karnataka state: the effect of death undercounting

We now present our analysis of Karnataka state viewed as an aggregate of all its districts. Karnataka has both urban and rural districts. We will assume that a suitable estimate of undercounting at the level of the entire state can be obtained through a population average of the undercounting at the level of its individual districts. Accounting for the fraction of the total population associated to each district, multiplying them by the district undercounting factor *U*(*D*) and summing these values gives us a population averaged death undercounting for Karnataka to be 2.2.

In this section we analyze the Karnataka infection curve in two ways. In the first, we assume no death undercounting. In the second we multiply the reported deaths by 2.2 to account for state-wide undercounting. For the calculations, the population data for Karnataka is sourced from the projections for the period 2020-2021, as provided by the report issued by Directorate of Economics and Statistics, Bangalore [100].

#### 2.3.1 Karnataka: No death undercounting

In Figure 9 we provide model results for Karnataka state data assuming deaths have been counted accurately. The posterior distributions and parameter constraints are provided in the SI, Section 3, Figure 7 and Table 6). Both infected and death numbers are well described by the model. The state-wide data has a single peak in both infected and deaths, indicating that the many variable peaks present in the individual districts are averaged out to yield the relatively simple structure shown in the figure. The numbers for those totally infected ranges between 15-45% at the 2*σ* level by mid February. We see that the strong urban to rural gradient in seropositivity, once averaged across the state, yields infected fractions that are below those in the urban regions. The bias multiplier follows results obtained from the city of Bengaluru.

**Table 6.** Mean and 95% bounds on the parameters. These values correspond to the posteriors plotted in Figure 12.

| Parameter | 95% limits |
| --- | --- |
| $E_{initial}$ | $6580^{+1000}_{-900}$ (people initially exposed) |
| $\beta_1$ | $0.0755^{+0.0020}_{-0.0021}$ |
| $\beta_2$ | $0.1170^{+0.0071}_{-0.0073}$ |
| $\beta_3$ | $0.220^{+0.019}_{-0.019}$ |
| $\tau_1$ | $142.1^{+9.7}_{-8.7}$ (days) |
| $Node_1$ | $153.9^{+4.2}_{-4.2}$ (day since the onset of simulation) |
| $Node_2$ | $234.0^{+3.3}_{-3.4}$ (day since the onset of simulation) |
| $\Delta_{IFR}$ | $62^{+10}_{-10}$ (days) |
| $D_{IFR}$ | $108.4^{+7.5}_{-7.0}$ (days) |
| $b$ | $> 194$ (multiplier) |
| $\Delta_b$ | $245^{+16}_{-15}$ (days) |
| $\sigma_1$ | $0.365^{+0.031}_{-0.028}$ |
| $\sigma_2$ | $0.237^{+0.021}_{-0.019}$ |

The IFR shows a decreasing trend after lockdown with a final range of IFR between 0.05-0.12%, consistent with estimates from the serological surveys [99]. These surveys find an overall adjusted total prevalence of 27.7% (95%CI 26.1–29.3); we obtain about 25% at the mid-line of the 95% confidence interval in mid-September. The case-to-infection ratio was estimated to be 1:40 while the infection fatality rate was 0.05%; our results are consistent with both observations. The *R*(*t*) band shows an increase after the lockdown till August 2020. After that *R*(*t*) decreases till December 2020 and stays below 1 till February 2021. In this case, bounds on cumulative infections and deaths are plotted with data in SI (Ref. Section 3, Figure 13).

#### 2.3.2 Karnataka: Incorporating death undercounting

Figure 10 shows our model predictions for the Karnataka state data where the death timeseries is uniformly scaled with multiplier of 2.2. The related shifts in the posterior distributions and parameter constraints can be seen in the SI : Section 3, Figure 7 and Table 6). The daily infection and the scaled daily death report fits are similar to Figure 9. However compared to the discussion that assumed no death multiplier predictions for the fraction of those infected ranges between 30-50% consistent with Table 5. A larger death undercounting thus leads to a higher estimates of actual infection within the population. Correspondingly, the bias multiplier is also large.

With higher death numbers, the effective IFR decreases at a later stage (around September 2020), convering around February to the values without death undercounting. The effect of death undercounting is absorbed mainly by the bias multiplier and therefore is reflected in the actual infected numbers. We find that the *R*(*t*) is nearly identical to the analysis without death undercounting. Assuming death undercounting, our estimates for the cumulative infections and deaths are plotted alongside the data in SI Section 3, Figure 14.

### 2.4 India

To describe the data for India, we assume that the death undercounting factor that represents India as a whole is the same as that obtained for the state of Karnataka, a factor of 2.2. In doing this we are motivated by the observations that the urban-rural gradient can be substantial and that the division between urban and rural populations at the Karnataka state level and at the national level are not too different, at an approximate ratio of about 35:65.

The national data for COVID-19 infections shows a single peak structure around mid-September 2020, at a bit less than 98,000 cases. The daily numbers then began to reduce after that reaching a value of about 10,000 by January of 2021. Our model fits infection and death reports in three adaptive windows between March 1, 2020 and February 15, 2021. For our calculations, we use India’s population data from the projections reported in the 2019 report published by National Commission on Population [107].

We find the bias multipliers change from 120 to 20 in this time period, thus indicating that by the end of the first wave, infections were being underestimated by the reported cases through a factor of 20. This decrease is in accordance with the reported overall increase in testing during this period. At the 2*σ* level, by the middle of February when the second wave began, we find that between 20-50% (95% CI) of Indian population was infected, with a point estimate of about 40%. This is a more conservative estimate than made by others. However, we believe it is consistent with the third ICMR serosurvey across December and January as well as with the slow pace of case accumulation over January and February.

The IFR band is consistent with a global India IFR of about 0.1% (95%CI 0.05 - 0.15). The *R*(*t*) band has several interesting features corroborated with the independent measurement plotted in dots. Before the lockdown, the *R*(*t*) stays in the high range 2.5-3. During lockdown it reduced to 1.5 and thereafter to the vicinity of 1.2. It stayed around 1.2 till August, going below 1 around October, signaling a steady decrease in infected numbers. A slight upward blip in November 2020 is consistent with the festival season in the north and east of India. The *R*(*t*) then stayed below 1 through till February 2021. Assuming death undercounting, estimations for cumulative infections and deaths for India are plotted with data in SI (Ref. Section 3, Figure 14]5). We present the posterior distribution of model parameters for the national data analysis in Figure 12.

Our estimates indicate that at the beginning of March the mean exposed population in India was around 6000. Around 14 March the number of officially reported infections in India crossed 100, but we expect that a lot of infections must have escaped detection.

Three adaptive windows are defined between simulation onset (March beginning) to 154’th day from the onset (August 2, 2020); between August 2, 2020 to October 21, 2020 (234th day from the simulation onset) and October 22, 2020 to February 15, 2021 (simulation end). These 3 regions have three different associated infectivities *β*_1_ = 0.0755, *β*_2_ = 0.117 and *β*_3_ = 0.220. This is the base infectivity with respect to the simulation onset. This is suggestive of two phases in which increased relaxations of COVID-19-associated restrictions may have led to a rise of cases, one around the beginning of August and one around the end of October, although one should be careful not to overinterpret this data,

Note that the effective *β* is modified by the parameter *τ* which has a mean value of 142 days. Therefore while the *β* is increasing with different windows, the effective *β* is also increasing. The bias parameter is unbounded from above even with a conservative prior. However the time evolution of bias determined by *b* and Δ_*b*_ for national data indicates a steady decrease, reaching a bias of around 20 as plotted in Figure 11. The IFR shows a transition around mid July (108 day since the onset) with a width Δ_IFR_ of 2 months. *σ*_1_ and *σ*_2_ represents the noise in the infected and deaths data respectively. They are well constrained.

The triangle plot of posteriors reveal strong correlations between parameters. Given the time-series of infected persons, a fit to the initial data can be addressed either by having a large initial exposed and lower infectivity or by having a smaller exposed population and a higher infectivity. Such a negative correlation is seen in Figure 12 between *E*_*initial*_ and *β*_1_. Since smaller *τ*_1_ results in a faster decrease in infection by lowering the effective *β*, to fit the infection data, if *β*_*i*_ increases, *τ* must decrease to maintain a similar level of effective infectivity. These negative correlations are reflected in the contour plots as well. The slope of the infection curve is given by *β*; the *β*’s are thus positively correlated.

## 3 Discussion and Conclusions

This paper describes the application of an epidemiological compartmental model, INDSCI-SIM, to COVID-19 in India. We focused on describing the first wave, starting from the first case in India on January 30, 2020 and continuing till mid-February, 2021, when a second and more destructive wave was initiated. Our intent was to obtain reasonable estimates for the fraction of the Indian population that was infected prior to the onset of the second wave. A parallel aim was to arrive at credible estimates of the Infection Fatality ratio (IFR) and of the undercounting of both cases and deaths during the first wave.

Assuming that deaths were recorded accurately in one urban district of Karnataka, we chose an infection fatality ratio for which our predicted fraction of infections and the fraction obtained through serosurveys in early September 2020 were approximately equal. We then used this to determine the extent of death undercounting across other districts of Karnataka, both urban and rural, by examining consistency between the numbers of infected as estimated through serosurveys and the numbers of deaths that should have resulted given the IFR value. We found that some districts could have under-counted deaths by a factor of about 5. Rural districts, in general, tended to have lower levels of infection spread, likely associated to the fact that agricultural labour and a good fraction of the rural workforce work outdoors, where infection risk is decidedly less. We thus estimated the overall fraction of infected in Karnataka to lie in the interval 30%-50%. Then, factoring this death undercounting into our calculation, we switched to a more refined calculation in whicn the IFR itself was allowed to vary. Averaging over all districts weighted by their populations yielded a factor of 2.2 between total actual and reported deaths in Karnataka state. We used these results to estimate that between 20-70% of the population across these districts were infected by February 2020.

Our results for the cities of Mumbai, Delhi, Pune and Bangalore - we assume no death undercounting here - indicate that, by February 2021, almost 60-80% of the population were infected, consistent with serological surveys in these cities. We found that only one in 15-20 cases were detected overall [112]. The effective reproductive ratio in all cases settled to just below 1 between December 2020 and February 2021, a period where cases were declining across India.

Our analysis for India, incorporating the same level of death undercounting as for Karnataka state, estimates that a fraction of about 40% (CI: 20-50%) have been infected at the 95% confidence level by the end of the first wave. We believe that this estimate, distinctly on the lower side of estimates by others, may account for the speed at which the second wave has spread. It may also account for the fact that rural spread seems to have been far larger in the second wave as opposed to the first, although the plight of urban India has tended to attracted more attention.

Our IFR estimates, though broad, are consistent with an inferred IFR of about 0.1%, with a broad 95% CI of (0.05 - 0.15). These are, as mentioned earlier, on the lower side *vis a vis*. population-based estimates for LMICs in general. We caution here that the extent of death undercounting will impact any estimate of the IFR. There are, as yet, no completely credible estimates of the extent of undercounting in the first COVID-19 wave in India [50, 113, 114]. In principle, we could have repeated the district-wise analysis we performed for Karnataka state for all states in India, but would be faced with the same problem, that of finding a benchmark district where we could safely assume that deaths were being counted more-or-less accurately (or knowing the extent of undercounting precisely) and then adjusting the mortality figures of other districts accordingly. We expect that as more data is obtained through better proxies for COVID-19 deaths or through accurate estimations of all-cause mortality, we should be able to refine these calculations further with more precise input.

The results presented here should be valuable in determining the initial conditions for the second wave, especially since we compute the posterior probabilities for the parameters which enter our model. Given these, and additional epidemiological and clinical input, for example the fraction of reinfections upon the introduction of a new strain, the decay of humoral immunity, the persistence and extent of cellular immunity, a vaccination program that was being slowly ramped up at the time of onset of the second wave as well as other features of the Indian response across February and March of 2021, we can then begin to address the question of what led to the fast increase of cases in the second wave. Our methodological innovations include accounting for a time-dependent improvement in case identification as well as in treatment leading to lower mortality, all implemented within a fully Bayesian framework. Questions such as the issue of reinfections as a consequence of immune escape, of partial immunity from the ongoing vaccination program, of vaccine breakthroughs at a low rate and of the impacts of multiple lock-downs applied inhomogeneously throughout the country should factor into any further analysis of events after the end of the first wave of COVID-19 in India. These impact public health policy intimately, especially as they are relevant to both testing strategies and vaccination policies [115, 116]. It is here that we expect that well formulated and bench-marked models should be of substantial use, both to understand the past better as well as to project the future more accurately.

## 4 Codes developed and used

We developed a fast FORTRAN code `ELiXSIR` *– Extended, zone Linked IX-compartmental SIR model: a code to simulate COVID19 infection* as a solver for the model [117]. A version of `ELiXSIR` is available for download in https://gitlab.com/dhirajhazra/eSIR_INDIA. The code is designed for arbitrary number of age groups and regions. This code is slightly modified for accommodate all parameters in the adaptive parametrization discussed in the text and compare with the data.

We acknowledge the use of `PolyChord` through the publicly available code `CosmoChord` https://github.com/williamjameshandley/CosmoChord. `ELiXSIR` is fused with `CosmoChord` for the purpose of data analysis and post processing. A detailed discussion of our analysis in SI (Ref. SI: Section 6).

## Data Availability

We developed a fast FORTRAN code (ELiXSIR) - Extended, zone Linked IX-compartmental SIR model: a code to simulate COVID19 infection} as a solver for the model. A version of ELiXSIR is available for download in https://gitlab.com/dhirajhazra/eSIR_INDIA. The code is designed for arbitrary numbers of age groups and regions. We acknowledge the use of PolyChord through the publicly available code CosmoChord: https://github.com/williamjameshandley/CosmoChord. ELiXSIR is fused with CosmoChord for the purpose of data analysis and post processing.

## Supporting Information (SI)

### SI: Section 1: Computation of reproduction number *R*_0_

We provide the calculation of the basic reproductive ratio corresponding to the age-stratified model, assuming uniform infectivity and values of *ϵ*= 1, using a next-generation-matrix method.

### SI: Section 2: Contact matrices between stratified age-groups

We describe how the contact matrices of Ref. [77] can be specified for our use here, combining contact matrices with a 5-year resolution into a more coarse-grained description.

Figure 1 : The coarse-grained contact matrices for India during (top) and without lockdown (bottom)

### SI: Section 3: Posterior distributions and correlations between parameters

We discuss the posterior distributions that emerge from our calculations, with figures and tables corresponding to our analysis for multiple cities, for Karnataka state and for India-wide numbers.

**Figure 2: Bengaluru Urban analysis** Marginalized posteriors of the parameters

**Table 1: Bengaluru Urban analysis** Constraints on parameters

**Figure 3: Chennai analysis** Marginalized posteriors of the parameters

**Table 2: Chennai analysis** Constraints on parameters

**Figure 4: Delhi analysis** Marginalized posteriors of the parameters

**Table 3: Delhi analysis** Constraints on parameters

**Figure 5: Mumbai analysis** Marginalized posteriors of the parameters

**Table 4: Mumbai analysis** Constraints on parameters

**Figure 5: Pune analysis** Marginalized posteriors of the parameters

**Table 5: Pune analysis** Constraints on parameters

**Figure 7: Karnataka analysis** Marginalized posteriors of the parameters

**Table 6: Karnataka analysis** Constraints on parameters

### SI: Section 4

Plots for cumulative infection and deaths

**Figure 8: Bengaluru Urban:** Bounds on cumulative infection and deaths plotted with reported data

**Figure 9: Chennai** Bounds on cumulative infection and deaths plotted with reported data

**Figure 10: Delhi** Bounds on cumulative infection and deaths plotted with reported data

**Figure 11: Mumbai** Bounds on cumulative infection and deaths plotted with reported data

**Figure 12: Pune** Bounds on cumulative infection and deaths plotted with reported data

**Figure 13: Karnataka - no death undercounting** Bounds on cumulative infection and deaths plotted with reported data. Here no death undercounting is assumed.

**Figure 14: Karnataka - incorporating death undercounting** Bounds on cumulative infection and deaths plotted with reported data. We assumed a multiplying factor of 2.2 (estimated using the Serosurvey data) for death undercounting.

**Figure 15: India** Bounds on cumulative infection and deaths plotted with reported data. We assumed a multiplying factor of 2.2 (estimated using the Serosurvey data) for death undercounting.

### SI: Section 5

Brief discussion on Nested Sampling. Here we very briefly outline how `PolyChord` works as a sampler.

### SI: Section 6

Flowchart of our analysis. Here we present the logical flow of our analysis. We discuss given a data and a model how our analysis pipeline works to obtain the constraints.

**Figure 16: Flowchart** Schematic diagram of our analysis.

## Acknowledgements

The numerical simulations discussed in the paper have been performed at the Institute of Mathematical Science’s High Performance Computing facility Nandadevi. DKH would like to thank Xingang Chen, Will Handley and Joshua Speagle for important discussions. BSP and SMS would like to thank Mihir Arjunwadkar for early discussions. GIM would like to thank Siva Athreya, Sandeep Krishna, Brian Wahl, Philip Cherian, Vaibhhav Sinha, Giridhara Babu, Bhramar Mukherjee, Gagandeep Kang, L S Shashidhara, Shahid Jameel, Kayla Laserson and Harish Iyer for many related discussions. SMS acknowledges funding from the DST-INSPIRE Faculty Fellowship (DST/INSPIRE/04/2018/002664) by DST India. GIM thanks the Department of Biological Science, TIFR (Mumbai) for an Adjunct Professorship and acknowledges support of the Bill and Melinda Gates Foundation, Grant No: R/BMG/PHY/GMN/20

