## Supplemental File for "The INDSCI-SIM model for COVID-19 in India"

### Supplementary materials for article: The INDSCI-SIM model for COVID-19 in India

June 2, 2021

#### Contents

**1 Computation of reproduction number $R_0$** **2**

**2 Contact matrices between stratified age-groups** **3**

**3 Posterior distributions and correlations between parameters** **3**

**4 Plots for cumulative infection and deaths** **3**

**5 Brief discussion on Nested Sampling** **3**

**6 Flowchart of our analysis** **5**

### 1 Computation of reproduction number $R_0$

Assuming uniform contacts across all age groups using the infectivity parameter $\beta$ and efficiency parameters $\epsilon_i$, the $\mathcal{F}$ matrix [1, 2] can be written as:

$$\mathcal{F} = \begin{pmatrix} 0 & \beta\epsilon_a(t) & \beta\epsilon_p(t) & \beta\epsilon_m(t) & \beta\epsilon_s(t) \\ 0 & 0 & 0 & 0 & 0 \\ 0 & 0 & 0 & 0 & 0 \\ 0 & 0 & 0 & 0 & 0 \\ 0 & 0 & 0 & 0 & 0 \end{pmatrix} \tag{1}$$

Here $a, p, m, s$ correspond to asymptomatic, pre-symptomatic, mild and severe compartments respectively.
And the $\mathcal{V}$ matrix is obtained as,

$$\mathcal{V} = \begin{pmatrix} \gamma & 0 & 0 & 0 & 0 \\ -\alpha\gamma & \lambda_a & 0 & 0 & 0 \\ -1(1-\alpha)\gamma & 0 & \lambda_p & 0 & 0 \\ 0 & 0 & -\mu\lambda_p & \lambda_m & 0 \\ 0 & 0 & -(1-\mu)\lambda_p & 0 & \lambda_s \end{pmatrix} \tag{2}$$

Here we have used the fractions and rates as defined in the main article.

$$\mathcal{V}^{-1} = \begin{pmatrix} \frac{1}{\gamma} & 0 & 0 & 0 & 0 \\ \frac{\alpha}{\lambda_a} & \frac{1}{\lambda_a} & 0 & 0 & 0 \\ \frac{1-\alpha}{\lambda_p} & 0 & \frac{1}{\lambda_p} & 0 & 0 \\ \frac{\mu-\alpha\mu}{\lambda_m} & 0 & \frac{\mu}{\lambda_m} & \frac{1}{\lambda_m} & 0 \\ \frac{(\alpha-1)(\mu-1)}{\lambda_s} & 0 & \frac{1-\mu}{\lambda_s} & 0 & \frac{1}{\lambda_s} \end{pmatrix} \tag{3}$$

The next generation matrix:

$$\mathcal{F}\mathcal{V}^{-1} = \begin{pmatrix} \beta\left(\frac{\alpha\epsilon_a(t)}{\lambda_a} - \frac{(\alpha-1)\mu\epsilon_m(t)}{\lambda_m} - \frac{(\alpha-1)\epsilon_p(t)}{\lambda_p} + \frac{(\alpha-1)(\mu-1)\epsilon_s(t)}{\lambda_s}\right) & \frac{\beta\epsilon_a(t)}{\lambda_a} & \beta\left(\frac{\mu\epsilon_m(t)}{\lambda_m} + \frac{\epsilon_p(t)}{\lambda_p} - \frac{(\mu-1)\epsilon_s(t)}{\lambda_s}\right) & \frac{\beta\epsilon_m(t)}{\lambda_m} & \frac{\beta\epsilon_s(t)}{\lambda_s} \\ 0 & 0 & 0 & 0 & 0 \\ 0 & 0 & 0 & 0 & 0 \\ 0 & 0 & 0 & 0 & 0 \\ 0 & 0 & 0 & 0 & 0 \end{pmatrix} \tag{4}$$

The dominant eigenvalue of the next generation matrix is $R_0$.

$$R_0 = \beta\left(\frac{\alpha\epsilon_a(t)}{\lambda_a} + (\alpha-1)\left(-\frac{\mu\epsilon_m(t)}{\lambda_m} - \frac{\epsilon_p(t)}{\lambda_p} + \frac{(\mu-1)\epsilon_s(t)}{\lambda_s}\right)\right) \tag{5}$$

Since INDSCI-SIM model is age stratified and uses contact matrices, the $\mathcal{F}$ and $\mathcal{V}$ matrices are generalized according to the model. Given $N$ age groups and $M$ equations determining the Exposed and infected compartments we will have $\mathcal{F}$ and $\mathcal{V}$ matrices of order $NM \times NM$. In our model we have 9 age compartments and 5 equations for exposed and 4 infected compartments. Therefore the $\mathcal{F}$ and $\mathcal{V}$ are of order $45 \times 45$.

The modified $\mathcal{F}$ matrix can be written as:

$$\mathcal{F} = \begin{pmatrix} [0] & [\mathcal{B}_i(t)] & \dots \\ \vdots & \ddots & \\ [0] & \dots & [0] \end{pmatrix} \tag{6}$$

Here $[0]$ represents $N \times N$ matrix with all entries as 0. $[\mathcal{B}_i(t)]$ represents the time dependent effective infectivity matrix (order $N \times N$) for different infected compartments and is defined as follows:

$$[\mathcal{B}_i(t)] = \epsilon_i\tilde{\beta}\exp[-t/\tau_i][C], \tag{7}$$

where the $m, n$'th element of the matix $[C]$ are defined with the contact matrix $C_{mn}$ and the population fraction $f_m$ as $C_{mn} f_m / f_n$. Here $\tau_i$ parameter represents the characteristic time-scale describing the increased effectiveness of non-pharmaceutical interventions, as introduced in the main paper. Note that $\beta$ in the unifrom contact case and $\tilde{\beta}$ in the differential contacts are different. In the uniform contact $\beta$ represents the infectivity averaged over all contacts while in this case $\tilde{\beta}$ represents the infectivity per contact. Note that the contact matrix changes during lockdown and therefore changes the effective infectivity matrix.

Generalized $\mathcal{V}$ matrix is now denoted as,

$$\mathcal{V} = \begin{pmatrix} [\Gamma] & [0] & [0] & [0] & [0] \\ -[A\Gamma] & [\Lambda_a] & [0] & [0] & [0] \\ -[(\mathbb{1} - A)\Gamma] & [0] & [\Lambda_p] & [0] & [0] \\ [0] & [0] & -[M\Lambda_p] & [\Lambda_m] & [0] \\ [0] & [0] & -[(\mathbb{1} - M)\Lambda_p] & [0] & [\Lambda_s] \end{pmatrix} \tag{8}$$

In the above $NM \times NM$ matrix each entry in square brackets represents a $N \times N$ matrix. In our model $M = 5$. For models with more infected compartments, $M$ will increase for both $\mathcal{F}$ and $\mathcal{V}$ matrices. $\Gamma$, $\Lambda_i$'s represent diagonal matrices with the transition rates as the diagonal terms. In our case, we have assumed the rates of transfer between compartments are same for all age groups. In a more general model, where the rates are different, the diagonal entries will change. The asymptomatic fractions and mild fractions are also represented by diagonal matrices, $A$ and $M$ respectively. Note that our fractions are age dependent and therefore, the diagonal terms contain the fractions for 9 age groups.

With the $\mathcal{F}$ and $\mathcal{V}$ matrices prepared, we follow the same procedure as in the uniform contact case and the dominated eigenvalue of $\mathcal{F}\mathcal{V}^{-1}$ is estimated as $R_0$.

At any point in time, while the basic reproduction number $R_0$ can be obtained using the relations above, the effective reproduction number $R(t)$ takes into account the susceptible population remaining at time $t$ *w.r.t.* the initial susceptible population. Thus, $R(t) = S(t = t)R_0/S(t = 0)$ where the susceptible population at $t = 0$ and $t = t$ are given by $S(t = 0)$ and $S(t = 0)$ respectively.

### 2 Contact matrices between stratified age-groups

Mixing between different age groups is determined by age-specific contact matrices. To compute these, we use contact matrices provided by [3], estimated for the Indian population, for 16 age groups ranging between 0-80 years, tabulated at 5 year intervals.

We reduce the matrix to further coarse-grained age brackets as follows: Let the number of contacts of the $i$-th individual in the age category $j$ with any individual belonging to the age category $k$ be $C_{kj}(i)$. The mean number of contacts for age category $j$ with age category $k$ is thus $\bar{C}_{kj} = \sum_{i=1}^{N_j} C_{kj}(i)/N_j$, where $N_j$ is the population size in age category $j$. Suppose now that $k$ is subdivided into 2 finer categories $k_1$ and $k_2$, and we are given the entries $\bar{C}_{k_1k_1}$, $\bar{C}_{k_1k_2}$, $\bar{C}_{k_2k_1}$ and $\bar{C}_{k_2k_2}$, as well as, $\bar{C}_{jk_1}$, $\bar{C}_{jk_2}$, $\bar{C}_{k_1j}$ and $\bar{C}_{k_2j}$. We then have

$$C_{kk} = [N_{k_1}\bar{C}_{k_1k_1} + N_{k_1}\bar{C}_{k_2k_1} + N_{k_2}\bar{C}_{k_1k_2} + N_{k_2}\bar{C}_{k_2k_2}]/(N_{k_1} + N_{k_2}) \tag{9}$$

with the off diagonal terms defined as $C_{jk} = [N_{k_1}\bar{C}_{jk_1} + N_{k_2}\bar{C}_{jk_2}]/(N_{k_1} + N_{k_2})$, and $C_{kj} = \bar{C}_{k_1j} + \bar{C}_{k_2j}$. We use this reduced matrix $[C]$ in our analysis. We plot the contact matrices in Figure 1.

### 3 Posterior distributions and correlations between parameters

Using getdist we compute the posterior distributions of the parameters for different regions and they are presented in triangle plots. We also tabulate the $2\sigma$ bounds on the parameters in the following tables.

### 4 Plots for cumulative infection and deaths

The timeseries of daily infection and deaths generate cumulative infection and deaths. In the following plots we plot the bounds on cumulative infection and deaths. Note that since we fit the daily infection and death reports, the error parameters correspond to those values and not the cumulative values.

### 5 Brief discussion on Nested Sampling

Since the posterior probabilities are expected to be multi-modal Markov Chain Monte Carlo can not be used. Therefore we use nested sampling with `PolyChord` that is effective for higher dimensional parameter spaces [4, 5]. This algorithm uses live

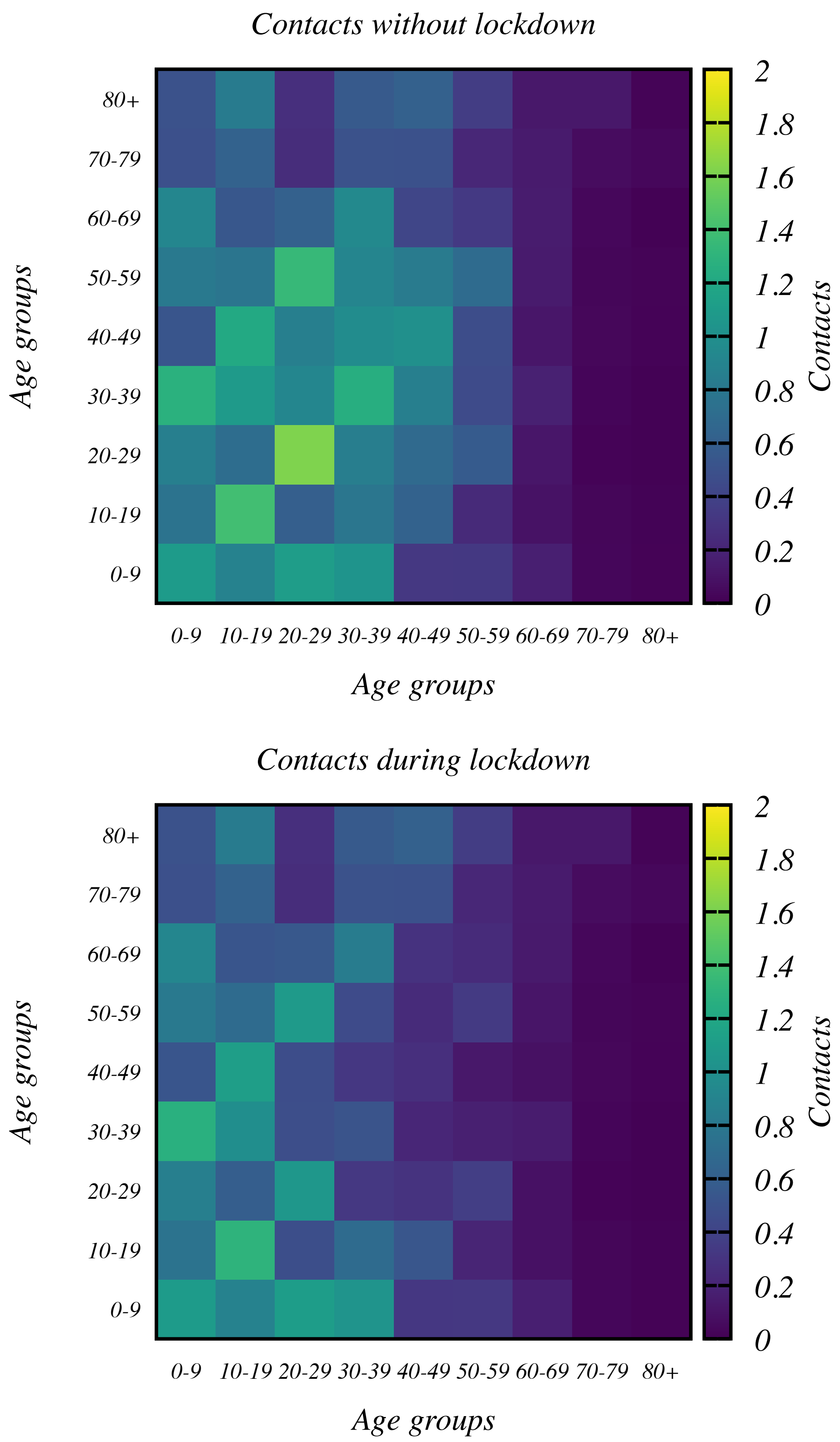

Figure 1: The coarse-grained contact matrices for India during (top) and without lockdown (bottom).

points that are updated in each iteration shrinking the n-dimensional parameter spaces. We use 500-1000 live points according to the dimension of the parameter space for the model. `Polychord` starts with the live points and the points are sequentially updated in each iteration. Point with the lowest live point is discarded (termed as dead point) and replaced by a new point

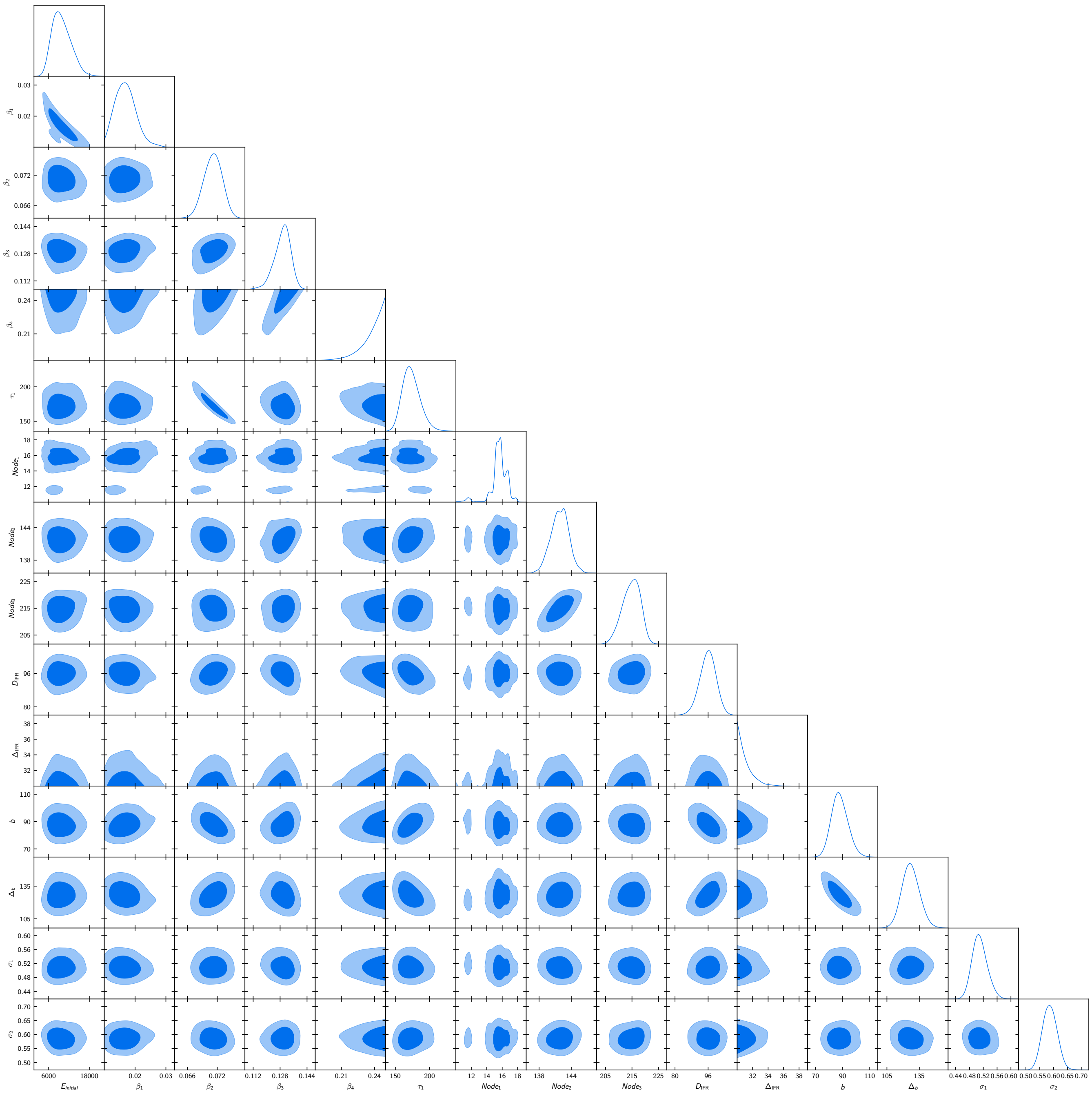

Figure 2: Marginalized posteriors of the parameters in the adaptive parametrization in the INDSCI-SIM model against the Bengaluru Urban data.

with a likelihood higher than that of the dead point. It can be shown [4] that the prior volume shrinks exponentially with each iteration. The evidence or the marginal likelihood is computed from the integral of likelihood and prior over the prior volume.

### 6 Flowchart of our analysis

In Figure 16 we plot the logical flow of the analysis. Centrally, we use 2 codes. For simulation, we have developed `ELiXSIR` – *Extended, zone Linked IX-compartmental SIR model: a code to simulate COVID19 infection* [6]. For sampling we use `PolyChord` [4], a widely used code for nested sampling. Both codes are available publicly.

Given a set of parameters we set up the system for `ELiXSIR`. In this set-up we provide the population, number of age groups and fraction of population in these age groups. We fix the fraction and rates of transition between compartments that are runtime constants. Lockdown dates are mentioned based on which the code switches from lockdown to unlock modes

| Parameter | 95% limits |
|---|---|
| $E_{initial}$ | $9805^{+6000}_{-5000}$ |
| $\beta_1$ | $0.0170^{+0.0063}_{-0.0066}$ |
| $\beta_2$ | $0.0711^{+0.0034}_{-0.0036}$ |
| $\beta_3$ | $0.1294^{+0.0090}_{-0.0098}$ |
| $\beta_4$ | $> 0.218$ |
| $\tau_1$ | $173^{+30}_{-20}$ |
| $Node_1$ | $15.6^{+1.8}_{-3.9}$ |
| $Node_2$ | $141.9^{+3.1}_{-3.3}$ |
| $Node_3$ | $214.8^{+6.1}_{-6.8}$ |
| $\Delta_{IFR}$ | $< 33.1$ |
| $D_{IFR}$ | $95.9^{+7.4}_{-8.1}$ |
| $b$ | $88^{+10}_{-10}$ |
| $\Delta_b$ | $127^{+20}_{-20}$ |
| $\sigma_1$ | $0.510^{+0.046}_{-0.039}$ |
| $\sigma_2$ | $0.586^{+0.051}_{-0.049}$ |

Table 1: Bengaluru Urban: Constraints on parameters. Corresponding to Figure 2 the 95% constraints and bounds are provided.

| Parameter | 95% limits |
|---|---|
| $E_{initial}$ | $178^{+50}_{-50}$ |
| $\beta_1$ | $0.0688^{+0.0023}_{-0.0024}$ |
| $\beta_2$ | $0.0429^{+0.0027}_{-0.0025}$ |
| $\beta_3$ | $0.0671^{+0.0088}_{-0.0075}$ |
| $\beta_4$ | $0.091^{+0.016}_{-0.015}$ |
| $\beta_5$ | $< 0.158$ |
| $\tau_1$ | $> 516$ |
| $Node_1$ | $66.9^{+3.6}_{-3.6}$ |
| $Node_2$ | $148.6^{+6.6}_{-6.2}$ |
| $Node_3$ | $228.8^{+8.7}_{-8.7}$ |
| $Node_4$ | $305^{+29}_{-23}$ |
| $\Delta_{IFR}$ | $93^{+30}_{-20}$ |
| $D_{IFR}$ | $85^{+20}_{-20}$ |
| $b$ | $45^{+5}_{-5}$ |
| $\Delta_b$ | $> 379$ |
| $\sigma_1$ | $0.350^{+0.030}_{-0.029}$ |
| $\sigma_2$ | $0.474^{+0.040}_{-0.037}$ |

Table 2: Chennai: Constraints on parameters. Corresponding to Figure 3 the 95% constraints and bounds are provided.

switching contact matrices. We use the coarse grained contact matrices for each region using the population fractions within the age groups of base contact matrix and the coarse grained age groups [3].

After the initial set up, the priors, initial starting value of all parameter and widths are provided. Daily data of reported infection and deaths are supplied to the code with the dates.

The `CosmoChord` integrated with `ELiXSIR` is then run in several processors (MPI) in the cluster. The samples within the parameter volume are drawn and sent to `ELiXSIR` for the time-series. Daily infection and deaths are computed from the compartments. Bias evolution is generated according to the parametric form and the values of $b, \Delta_b$ from the samples. After scaling the theoretically obtained daily infection numbers by the bias, the prediction of daily infected is then compared with the reported daily infection. Predicted daily deaths however is directly compared with the daily deaths data. If the death undercounting factor is included in the analysis then the daily deaths prediction is compared with the death data scaled by the undercounting factor. During the runtime, live points are generated and updated sequentially that reduces the prior volume, as discussed in the last section.

After termination of the `PolyChord` sampling, we use getdist [7] to generate posterior distributions. The samples are then supplied to `ELiXSIR` directly to generate the bounds on the timeseries.

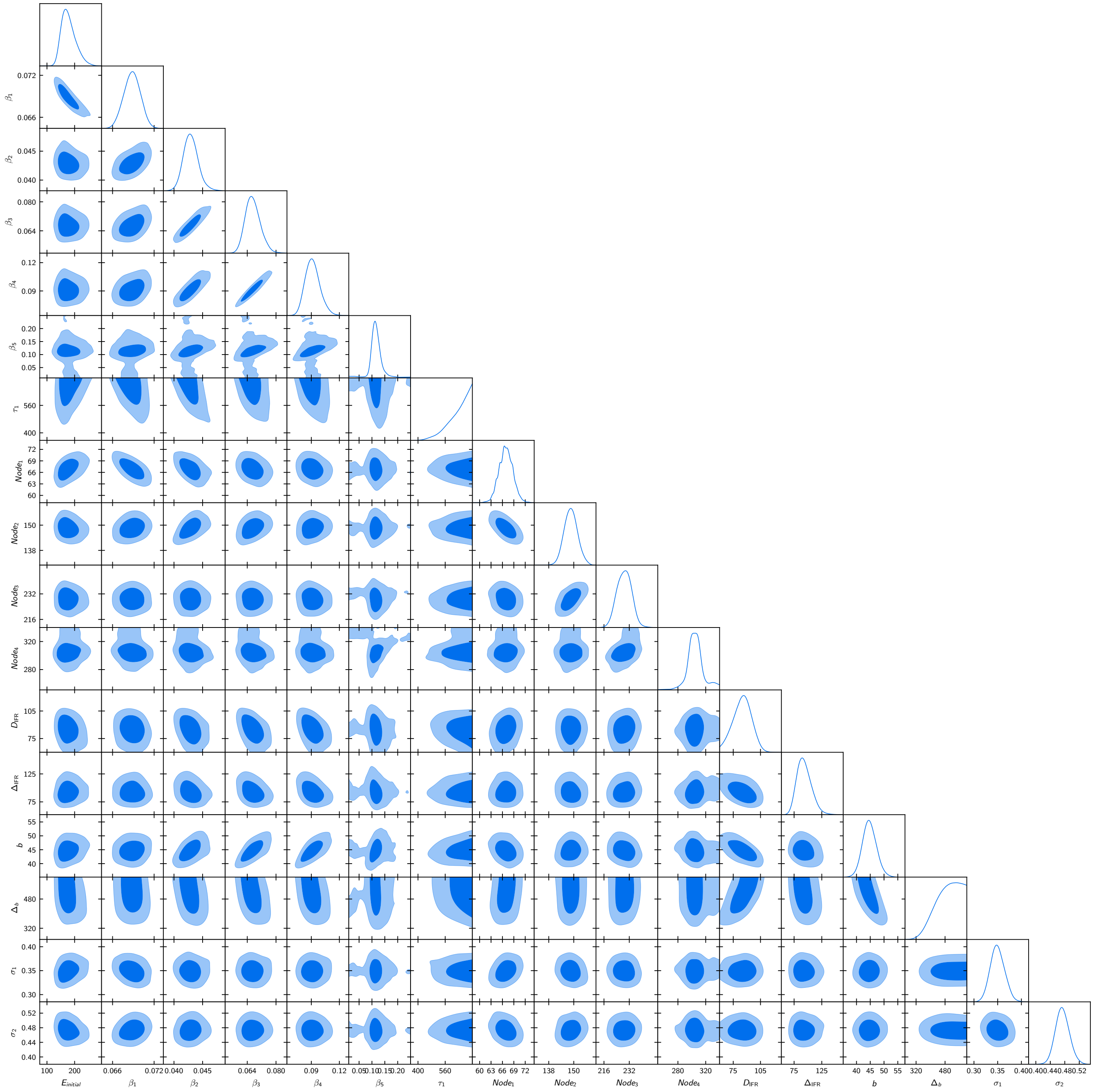

Figure 3: Marginalized posteriors of the parameters in the adaptive parametrization in the INDSCI-SIM model against the Chennai data.

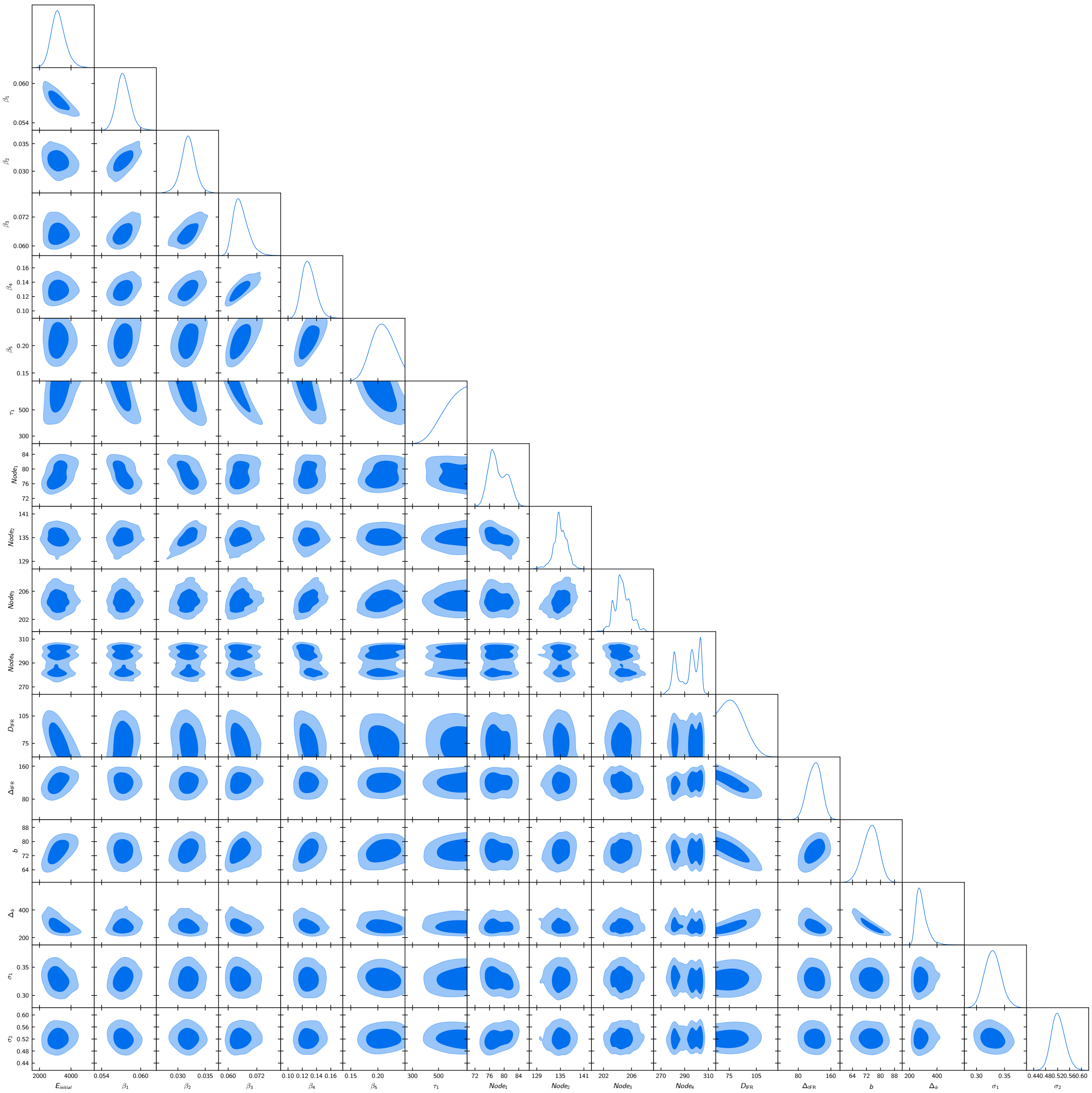

Figure 4: Marginalized posteriors of the parameters in the adaptive parametrization in the INDSCI-SIM model against the Delhi data.

[5] Handley W. fgivenx: Functional Posterior Plotter. The Journal of Open Source Software. 2018;3(28). doi:10.21105/joss.00849.

[6] Hazra DK. ELiXSIR – Extended, zone Linked IX-compartmental SIR model: a code to simulate COVID19 infection; 2021. Available from: `https://gitlab.com/dhirajhazra/eSIR_INDIA`.

[7] Lewis A. GetDist: a Python package for analysing Monte Carlo samples; 2019.

| Parameter | 95% limits |
|---|---|
| $E_{initial}$ | $3199^{+1000}_{-900}$ |
| $\beta_1$ | $0.0574^{+0.0021}_{-0.0021}$ |
| $\beta_2$ | $0.0319^{+0.0026}_{-0.0028}$ |
| $\beta_3$ | $0.0653^{+0.0066}_{-0.0057}$ |
| $\beta_4$ | $0.129^{+0.020}_{-0.018}$ |
| $\beta_5$ | $0.208^{+0.040}_{-0.034}$ |
| $\tau_1$ | $> 439$ |
| $Node_1$ | $78.3^{+4.7}_{-4.0}$ |
| $Node_2$ | $135.0^{+3.3}_{-3.8}$ |
| $Node_3$ | $204.7^{+2.3}_{-2.1}$ |
| $Node_4$ | $293^{+13}_{-15}$ |
| $\Delta_{IFR}$ | $120^{+30}_{-30}$ |
| $D_{IFR}$ | $< 102$ |
| $b$ | $74^{+8}_{-9}$ |
| $\Delta_b$ | $288^{+90}_{-70}$ |
| $\sigma_1$ | $0.329^{+0.030}_{-0.027}$ |
| $\sigma_2$ | $0.522^{+0.048}_{-0.043}$ |

Table 3: Delhi: Constraints on parameters. Corresponding to Figure 4 the 95% constraints and bounds are provided.

| Parameter | 95% limits |
|---|---|
| $E_{initial}$ | $674^{+200}_{-200}$ |
| $\beta_1$ | $0.0692^{+0.0035}_{-0.0030}$ |
| $\beta_2$ | $0.0440^{+0.0038}_{-0.0032}$ |
| $\beta_3$ | $0.085^{+0.013}_{-0.012}$ |
| $\beta_4$ | $0.150^{+0.033}_{-0.030}$ |
| $\tau_1$ | $> 388$ |
| $Node_1$ | $60.9^{+5.2}_{-5.7}$ |
| $Node_2$ | $153.5^{+2.9}_{-3.1}$ |
| $Node_3$ | $< 254$ |
| $\Delta_{IFR}$ | $53^{+10}_{-10}$ |
| $D_{IFR}$ | $< 94.3$ |
| $b$ | $156^{+15}_{-14}$ |
| $\Delta_b$ | $208^{+20}_{-20}$ |
| $\sigma_1$ | $0.470^{+0.040}_{-0.037}$ |
| $\sigma_2$ | $0.339^{+0.033}_{-0.029}$ |

Table 4: Mumbai: Constraints on parameters. Corresponding to Figure 5 the 95% constraints and bounds are provided.

| Parameter | 95% limits |
|---|---|
| $E_{initial}$ | $6670^{+1000}_{-1000}$ |
| $\beta_1$ | $0.0477^{+0.0013}_{-0.0012}$ |
| $\beta_2$ | $0.0735^{+0.0080}_{-0.0069}$ |
| $\beta_3$ | $0.167^{+0.033}_{-0.030}$ |
| $\beta_4$ | $0.214^{+0.058}_{-0.050}$ |
| $\tau_1$ | $> 540$ |
| $Node_1$ | $> 119$ |
| $Node_2$ | $195.2^{+3.5}_{-4.3}$ |
| $Node_3$ | $252^{+17}_{-12}$ |
| $\Delta_{IFR}$ | $< 55.9$ |
| $D_{IFR}$ | $138^{+12}_{-12}$ |
| $b$ | $49.9^{+4.4}_{-4.1}$ |
| $\Delta_b$ | $429^{+200}_{-100}$ |
| $\sigma_1$ | $0.393^{+0.034}_{-0.030}$ |
| $\sigma_2$ | $0.643^{+0.055}_{-0.051}$ |

Table 5: Pune: Constraints on parameters. Corresponding to Figure 6 the 95% constraints and bounds are provided.

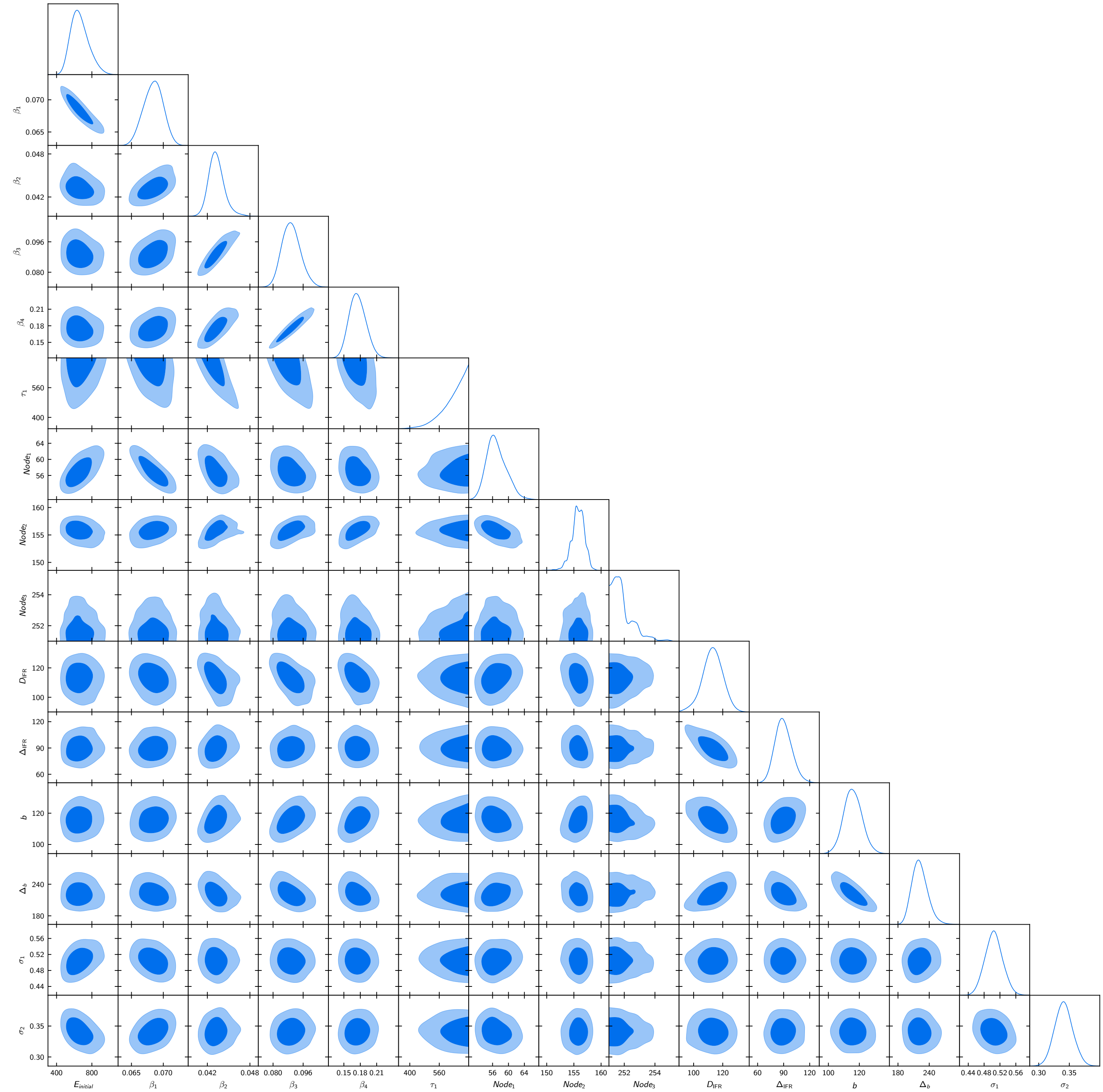

Figure 5: Marginalized posteriors of the parameters in the adaptive parametrization in the INDSCI-SIM model against the Mumbai data.

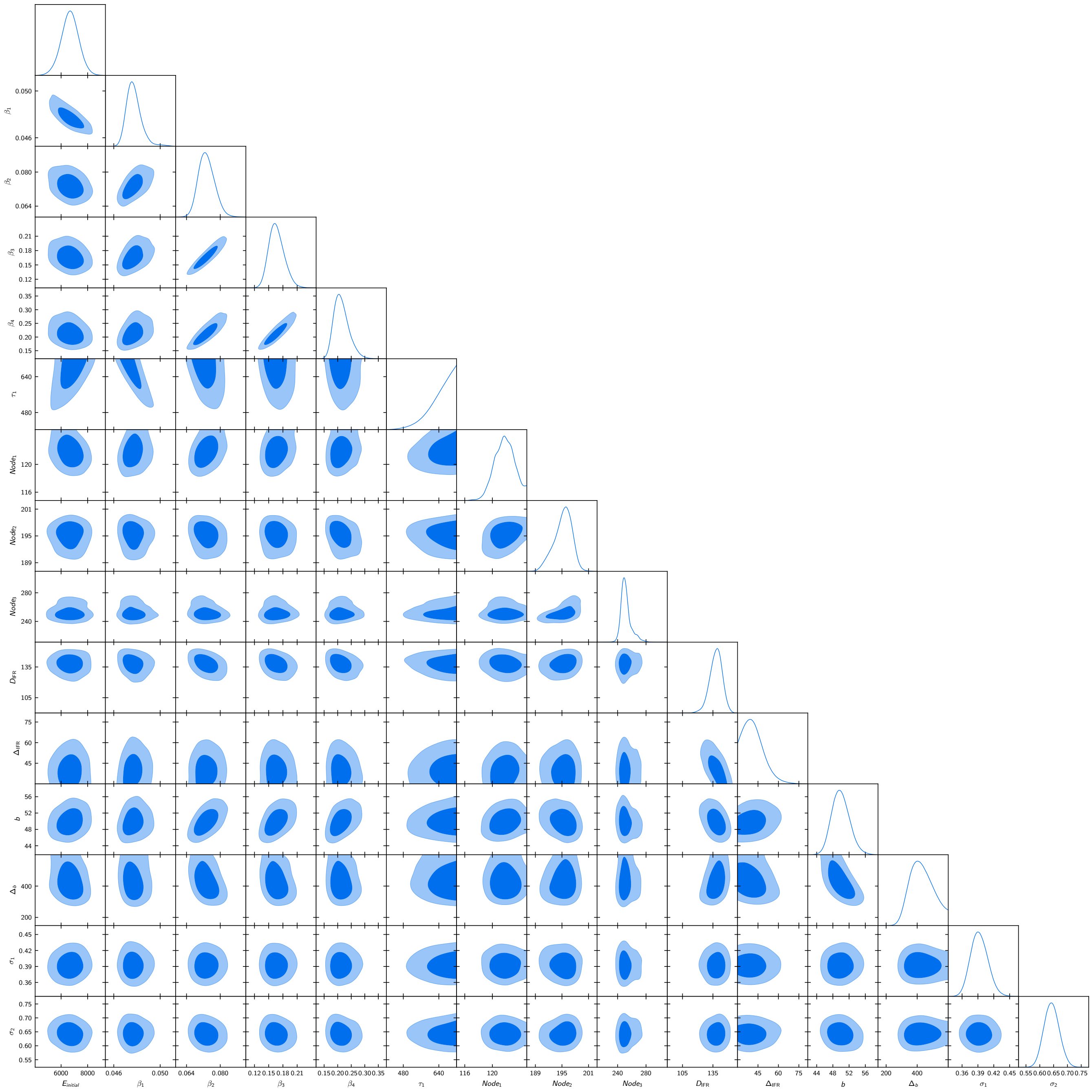

Figure 6: Marginalized posteriors of the parameters in the adaptive parametrization in the INDSCI-SIM model against the Pune data.

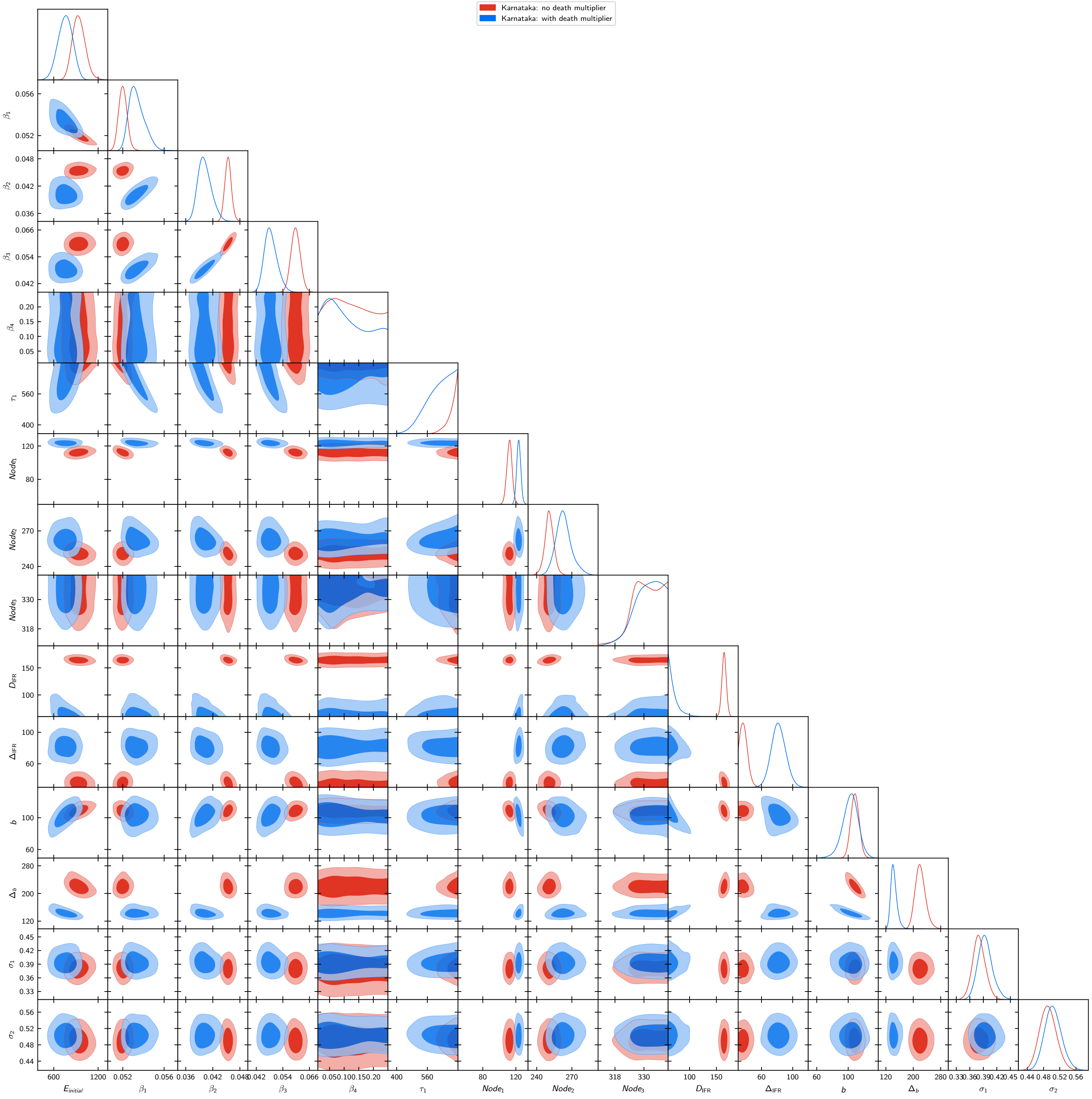

Figure 7: Marginalized posteriors of the parameters in the adaptive parametrization in the INDSCI-SIM model against the Karnataka data. We have plotted the constraints obtained assuming and without assuming death multiplier for undercounting.

| Parameter | 95% | limits |
|---|---|---|
| | Without multiplier | With multiplier |
| $E_{initial}$ | $757^{+200}_{-200}$ | $939^{+200}_{-200}$ |
| $\beta_1$ | $0.0534^{+0.0016}_{-0.0013}$ | $0.05201^{+0.00081}_{-0.00075}$ |
| $\beta_2$ | $0.0402^{+0.0031}_{-0.0027}$ | $0.0454^{+0.0015}_{-0.0014}$ |
| $\beta_3$ | $0.0486^{+0.0059}_{-0.0053}$ | $0.0597^{+0.0043}_{-0.0041}$ |
| $\beta_4$ | — | — |
| $\tau_1$ | $> 501$ | $> 631$ |
| $Node_1$ | $123.5^{+5.0}_{-4.8}$ | $112.3^{+6.3}_{-6.3}$ |
| $Node_2$ | $263^{+15}_{-12}$ | $250.8^{+7.8}_{-7.9}$ |
| $Node_3$ | $> 322$ | $> 321$ |
| $\Delta_{IFR}$ | $82^{+20}_{-20}$ | $< 46.4$ |
| $D_{IFR}$ | $< 92.2$ | $164.3^{+8.2}_{-8.1}$ |
| $b$ | $103^{+20}_{-20}$ | $109^{+10}_{-10}$ |
| $\Delta_b$ | $144^{+20}_{-20}$ | $221^{+30}_{-30}$ |
| $\sigma_1$ | $0.394^{+0.035}_{-0.029}$ | $0.380^{+0.029}_{-0.027}$ |
| $\sigma_2$ | $0.503^{+0.044}_{-0.039}$ | $0.491^{+0.042}_{-0.039}$ |

Table 6: Karnataka: Constraints on parameters. Corresponding to Figure 7 the 95% constraints and bounds are provided. Two columns on constraints represent the results when we use reported death data without and with undercounting multiplier respectively.

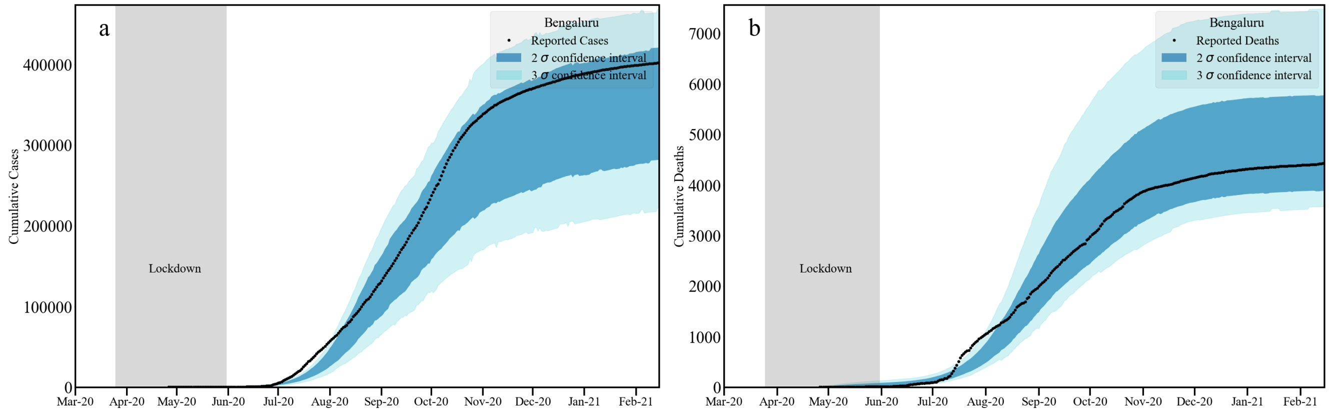

Figure 8: Bengaluru Urban: Bounds on cumulative infection [a: left] and deaths [b: right] from our analysis plotted with reported data.

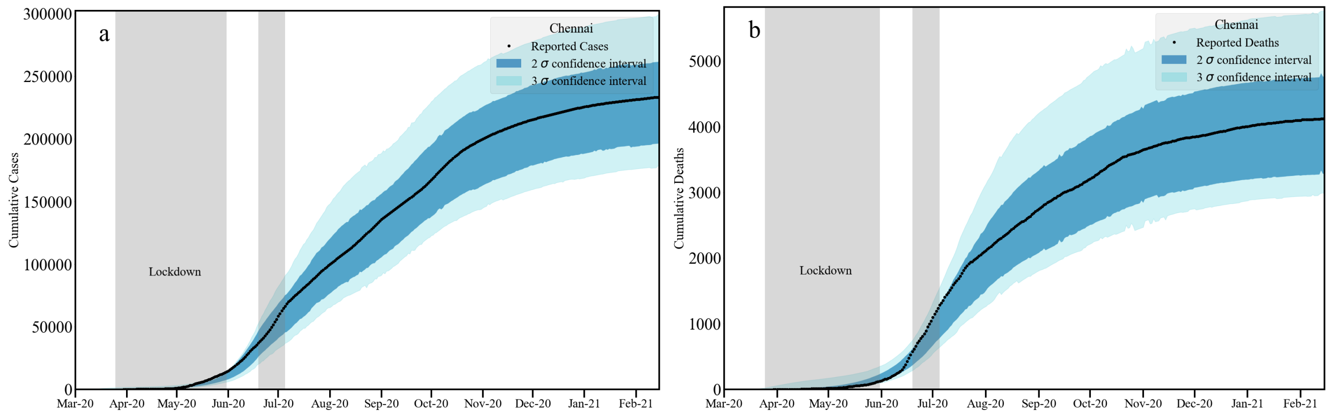

Figure 9: Chennai: Bounds on cumulative infection [a: left] and deaths [b: right] from our analysis plotted with reported data.

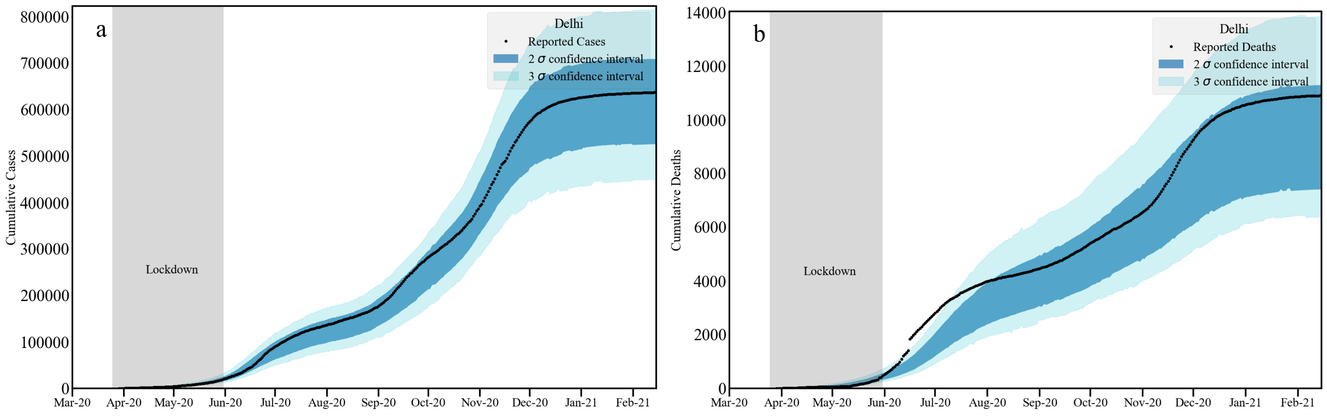

Figure 10: Delhi: Bounds on cumulative infection [a: left] and deaths [b: right] from our analysis plotted with reported data.

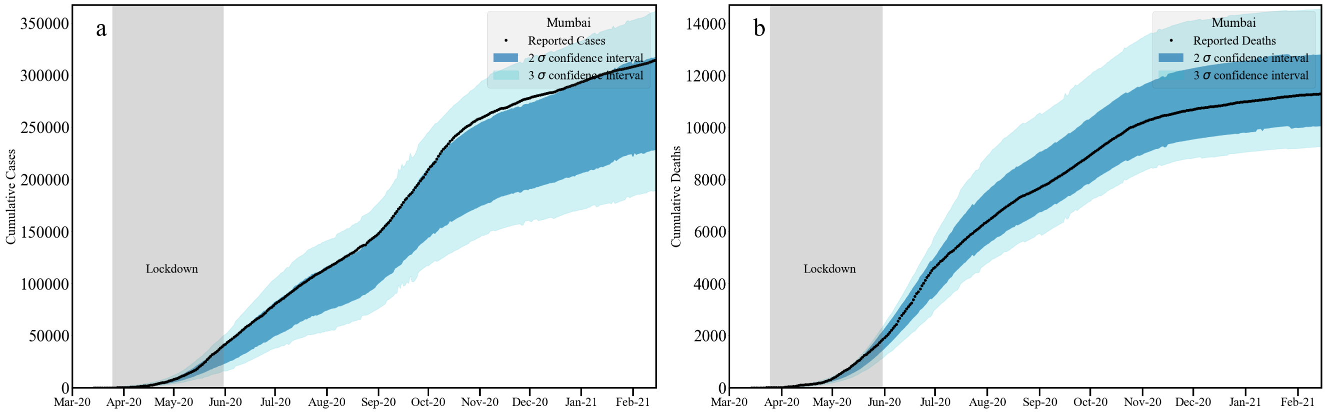

Figure 11: Mumbai: Bounds on cumulative infection [a: left] and deaths [b: right] from our analysis plotted with reported data.

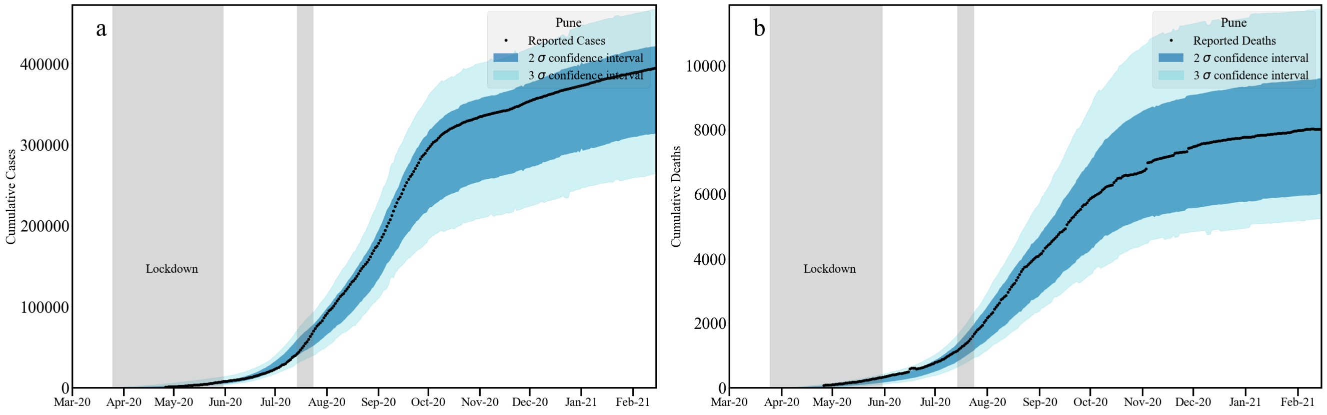

Figure 12: Pune: Bounds on cumulative infection [a: left] and deaths [b: right] from our analysis plotted with reported data.

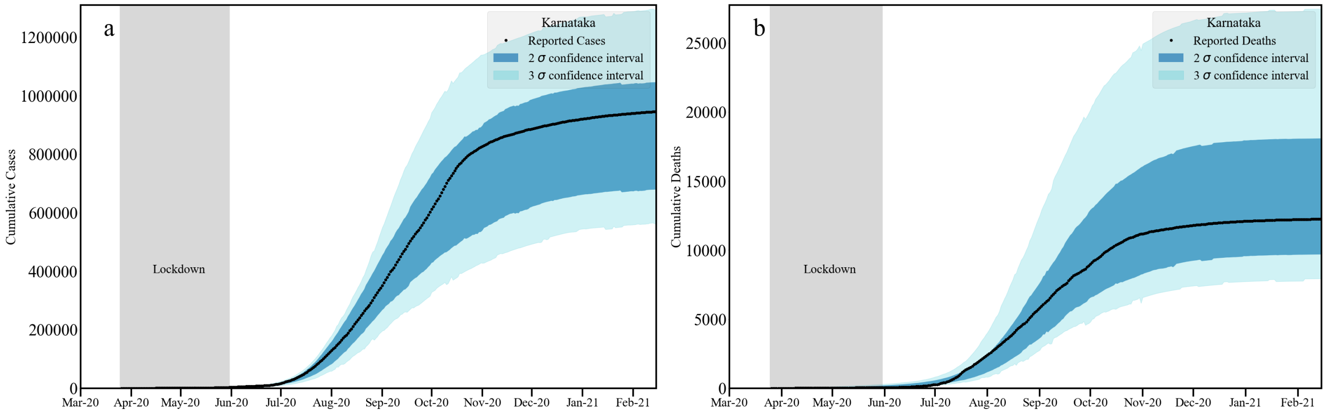

Figure 13: Karnataka: Bounds on cumulative infection [a: left] and deaths [b: right] from our analysis plotted with reported data.

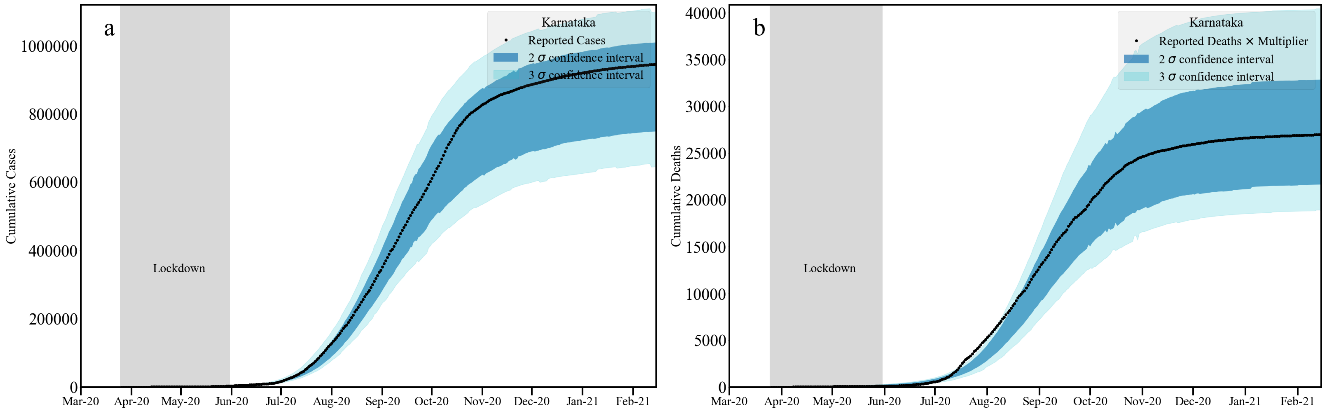

Figure 14: Karnataka: Bounds on cumulative infection [a: left] and deaths [b: right] from our analysis plotted with reported data. Note that here death multiplier 2.2 is used to take into account possible death undercounting.

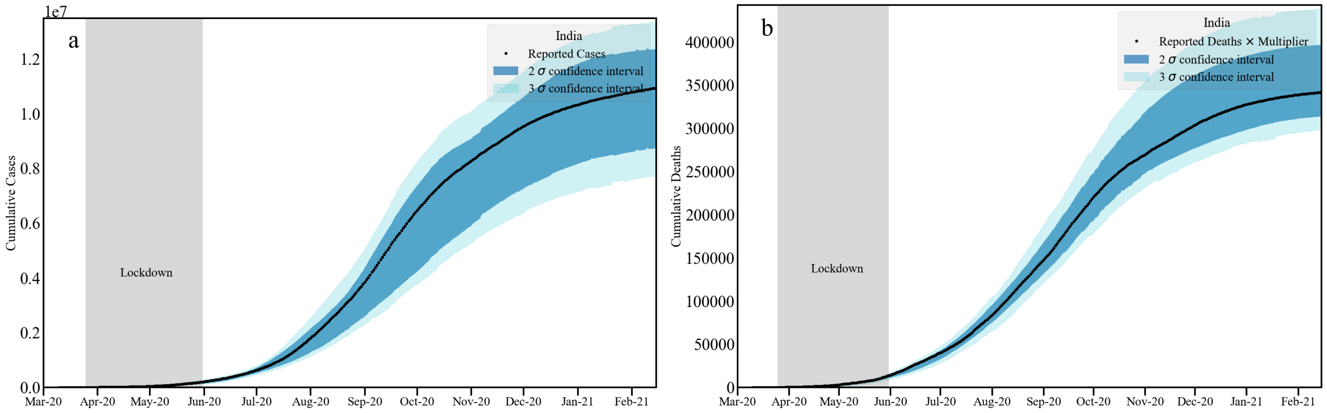

Figure 15: India: Bounds on cumulative infection [a: left] and deaths [b: right] from our analysis plotted with reported data. Note that here death multiplier 2.2 is used to take into account possible death undercounting.

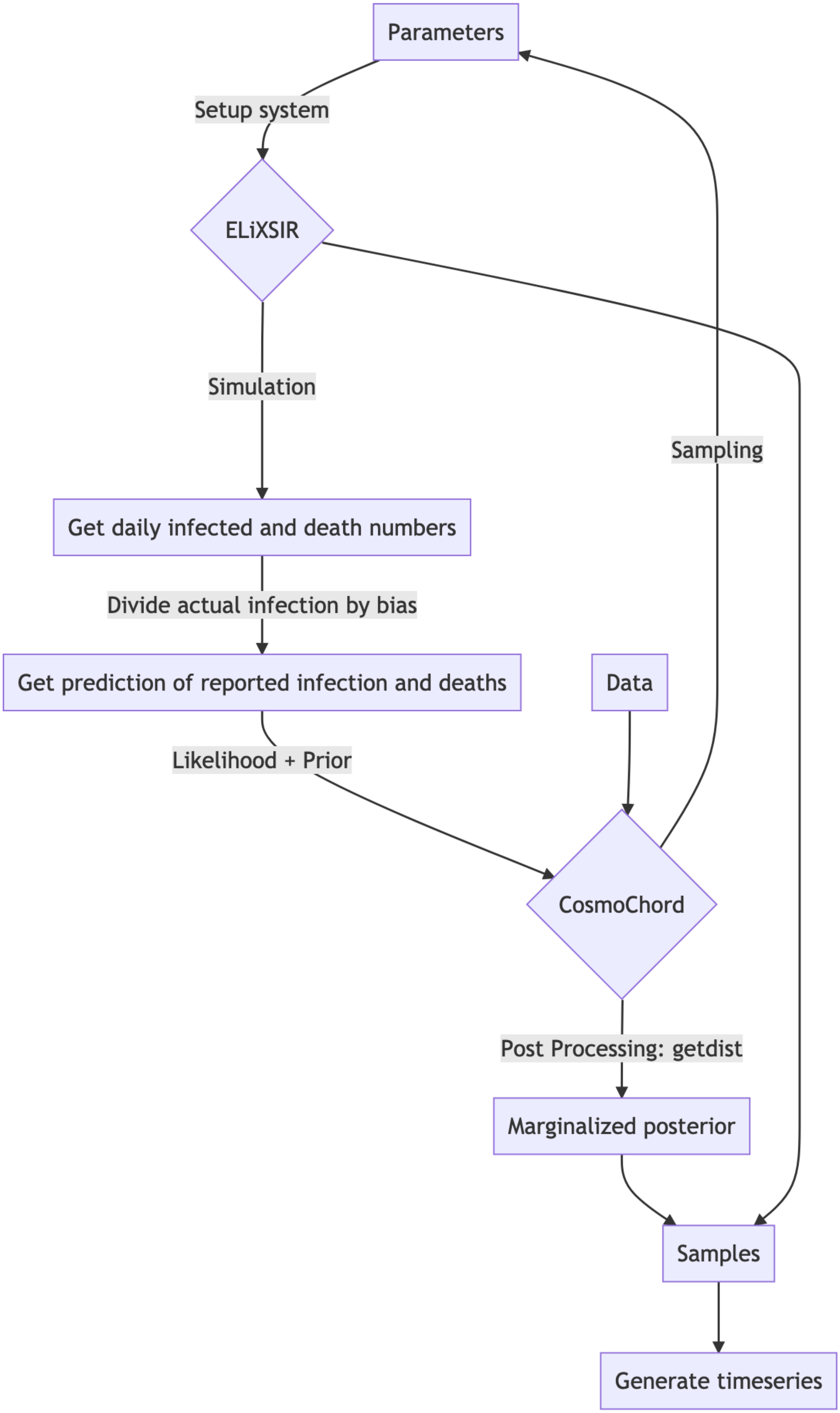

Figure 16: Schematic diagram of our analysis.
